# A Simulation Study of Sampling in Difficult Settings: Statistical Superiority of Little-Used method

**DOI:** 10.1101/2022.09.12.22279856

**Authors:** Harry S. Shannon, Patrick D. Emond, Benjamin M. Bolker, Román Viveros-Aguilera

**Affiliations:** McMaster University, Hamilton, Ontario, Canada, L8S 4K1

**Keywords:** sampling methods, Extended Program on Immunization (EPI), virtual populations, computer simulation, Global Positioning Systems (GPS), small area sampling

## Abstract

**Background:** Taking a representative sample to determine prevalence of variables such as disease is difficult when little is known about the target population. Several methods have been proposed that apply cluster sampling techniques to Primary Sampling Units (PSUs). The PSUs are typically towns or census tracts. Some methods are based on random walks within towns, e.g., the original World Health Organization’s Extended Program on Immunization (‘EPI’) surveys and variants, including sampling from four quadrants of each town (‘Quad’). Several major international surveys take random samples from small areas (‘SA’) such as census tracts. Another method uses satellite images and Global Positioning Systems to randomly sample within PSUs from squares in a superimposed grid (‘Square’). We used computer simulations to compare these sampling methods and simple random sampling within towns (‘SRS’) in virtual populations. SRS was our standard, even though it is impractical in low-information settings.

**Methods:** We constructed 50 virtual populations with varying characteristics, each comprising about a million people spread over 300 towns. The risk of disease for each person varied within and between towns. We created a binary exposure variable and allocated disease statuses to individuals assuming four relative risks (RRs) from exposure. We added three populations with equal risk of disease for every person in the population.

For each population, each of the sampling methods – EPI, Quad, SA, Square, and SRS - and each of three sample sizes per PSU (7, 15, and 30), we simulated 1000 samples. For each simulation we estimated the prevalence and RRs. We used the bias and variance of the estimates to calculate the Root Mean Squared Error (RMSE) of these estimates. We ranked the RMSEs of each method and computed the ratio of each method’s RMSE to that of SRS for each population. We computed the mean ranks and ratios across the 50 populations.

**Results:** Apart from SRS, Square had the lowest mean rank of RMSEs for all samples sizes when estimating prevalence. When estimating RR, Square had the lowest mean ranks for samples sizes of 15 and 30 per PSU; for n=7 per PSU, the Quad mean rank was the lowest.

The results for the mean ratios of RMSEs showed the same pattern; Square had the lowest values for all sample sizes when estimating prevalence and the lowest values for the two larger sample sizes when estimating RRs. Notably, when estimating prevalence, the ratios increased with sample size per PSU for SA, Quad, and EPI, suggesting those methods did not benefit as much from the larger sample sizes as would be expected from statistical theory.

**Conclusions:** Of several methods that are practical in an imperfectly known population, the Square method was mostly the best, especially for the larger sample sizes. The methods that sample within small areas (Quad, SA, and EPI) do not gain as much statistical benefit as expected from larger sample sizes per PSU, because of some clustering within the areas.

## Introduction

Health surveys in various parts of the world are conducted, e.g., to estimate prevalence of disease or immunization, or relative risk (RR) of disease given exposure to a putative hazard. Conducting these surveys can be challenging when relevant information on the population of interest, such as population size in different areas, is limited.

Various survey methods have been proposed for such low-information scenarios; some have been applied in the field. The World Health Organization (WHO) developed a random walk methodology to estimate immunization rates in young children as part of the Extended Program on Immunization (EPI) [World Health organization, 2005]. This approach selects 30 towns as Primary Sampling Units (PSUs) using probability proportional to size (PPS). To overcome the lack of a complete list of households, surveyors identify a central landmark in each town, choose a random direction, identify all households along that direction to the edge of the town, and randomly choose one as the starting household. Additional households are selected using a ‘nearest neighbor’ process until the required sample size is reached. We label this approach ‘EPI’. (The figures in the Appendix shows how it and other sampling methods are applied.)

EPI had limitations – in particular, the sampling probabilities are undetermined, making it difficult to construct adjusted, unbiased estimates from the survey results. Several authors have proposed modifications. For example, Bennett et al. [1994] suggested several approaches to ensure a wider geographic dispersion of the sample. One method divided the town into four quadrants and applied the EPI approach to select a quarter of the sample from each quadrant (‘Quad’). They proposed additional options: taking half the sample from the centre of the town and half from the edge; taking every third nearest house; and taking every fifth nearest house. Grais et al. [2007] recognized that EPI biases the starting house to be close to the center of the town and proposed an alternative method to identify the starting household. However, none of these changes allows the estimation of sampling probabilities.

Kolbe et al. [2010] made use of satellite images and Global Positioning Systems (GPS). They randomly chose GPS points within the survey area, drew circles around them on the images, numbered the buildings in the circle, and randomly chose one building from each circle (‘Circle’). Shannon et al. [2012] suggested a variant to avoid the overlap that can occur with circles: superimposing a grid of squares over the images of towns, randomly sampling several squares from each town, and randomly sampling a building from each square (‘Square’ – see Appendix for schematic figure). Ambiguities about buildings that overlap the edges of squares can be resolved by assigning buildings to a square based on the side on which the building falls (e.g. north/west vs. south/east).

Several surveys [e.g., MICS, 2019; Demographic and Health Surveys, n.d.] sample small areas (typically census Enumeration Areas), identify all households in those areas, and take random samples of the households in each area. The WHO has revised its EPI evaluation and uses this procedure of sampling small areas which we label ‘SA.’ [World Health Organization, 2018].

Some simulations have assessed whether the EPI method was ‘good enough,’ i.e., whether the biases and variances of the estimates were sufficiently small for the survey’s purposes [Henderson and Sundaresan, 1982; Lemeshow et al., 1985]. Bennett and colleagues [1994] concluded that the variants they suggested performed better than the EPI approach. Himelein et al. [2017] found that a random walk method performed poorly in estimating a continuous variable, household consumption.

We have conducted a simulation study whose aim was to compare the different sampling methods in estimating prevalence of a variable and RR of a disease given an exposure. We also examined how the performance of the methods depended on characteristics of the populations. We looked at sampling methods under ideal conditions and did not consider practical issues in surveys. Cutts et al. [2013] provide a discussion of these issues.

We investigated ten methods, including the variants of the EPI technique described by Bennett et al. [1994]. For clarity, we report only on simple random sampling and four other selected methods in this paper. We exclude most variants of the original EPI evaluation. The one we include (Quad) is the one that performed best in our simulations. Descriptions of all the methods and full results are in the supplementary information.

## Methods

Our broad approach was as follows:

- Create many virtual populations with known characteristics (parameters), including allocation of disease or vaccination status and an exposure and disease status for different relative risks (RRs) from that exposure.
- Simulate different sampling methods to take multiple samples from the populations.
- Estimate the prevalence of disease and the RRs from an exposure for each sample.
- For each method, combine the bias and variance of the samples to compute the Root Mean Square Error (RMSE) of the estimates.
- Compare the RMSEs for the different sampling methods both overall and in relation to the population characteristics

Henceforth, we label the binary outcome ‘disease.’

### Creating the virtual populations

The simulation program was written to be extremely flexible. A variety of parameters was chosen, as we attempted to mimic how those parameters might vary in real life. We varied parameters for the overall populations and for characteristics of towns, households, and individuals within populations. To consider a broad range of many different parameters we used a ‘Latin hypercube’ approach [Blower and Dowlatabadi, 1994], treating the parameters as measures that varied in small increments within a pre-specified range and ensuring unique combinations of the parameters. The technique is in effect a stochastic form of fractional factorial design that works well with large numbers of parameters.

For each simulated population, we randomly sampled one of the possible values for each parameter without replacement. For example, for the mean sizes of households the range was from 2 to 5, varying by units of 0.06. Since we created 50 populations with characteristics varying between and within the towns, we allowed 50 values for the parameters, and the Latin Hypercube approach ensured we used each of those values in exactly one simulated population. Other population parameters included the target disease prevalence, number of disease pockets per town (see below), and prevalence of exposure. The values used were chosen as for household size.

Each population created was distributed among cities, towns and villages (henceforth, simply ‘towns’) using a Pareto distribution. We created 300 towns, with population sizes between 400 and 250,000. Each town was geographically represented as a square, 10,000 by 10,000 units. Given a parameter value for a population, the actual value for a particular town was randomly chosen from a normal distribution centred at the population value with a small variance. Within each town, we divided the area into 100 smaller squares (a 10 × 10 grid) each 1,000 by 1,000 units, labelling the axes x and y. The values of x and y were used to determine the overall characteristics of people living in the sub-area. The first determination was the range in the density between the most and the least densely populated sub-areas. The density varied linearly with each of x and y, so that the minimum and maximum densities were at opposite corners of each town.

The households were placed within each square by randomly choosing spatial coordinates within the appropriate range. We did not require a minimum distance between households; any two households close together could be considered to be part of a multi-residence building. The number of people in a household was randomly determined, based on the hypercube value for the mean number per household, using a zero-truncated Poisson distribution. The first two people in the household were taken to be adults, and additional members were designated as children. Using the linear function that determined the population density, households were added until the sub-area had the predetermined number of people. Other characteristics, such as household income or age (adult vs. child), were determined using the other parameters. The supplementary information shows the parameters used in the simulations, and the ranges of possible values allowed.

We incorporated ‘pocketing’, the presence of small areas with particularly high prevalence, representing a local spread of infection. This was done by randomly identifying points that were the centres of pockets. The number of pockets per town was randomly chosen for each population. Each pocket added to the risk of disease for everyone in the town. The risk declined rapidly with distance from the centre of the pocket, so for most people the additional risk was minimal. The program allowed for different numbers of pockets and for differing functions and rates of decline with distance of the additional risk.

The allocation of disease status to each individual was based on the risk, which was in turn based on several factors. Once the background disease risk for a sub-area was determined, several additional factors played a role, including age and household income. Each person’s disease status was determined randomly based on the calculated binomial probability. The disease determination algorithm in Additional file 2 provides further detail. The random determination of disease status meant that the prevalence in a population differed from the target value that had been chosen.

We also incorporated bivariate relationships between variables representing an exposure and a disease. The likelihood of exposure varied across the population depending on the location of the household. We considered relative risks (RRs) of 1.0, 1.5, 2.0, and 3.0. To program these, we assigned a different disease for each RR; for Disease 1 we had RR=1.0, for Disease 2 we had RR=1.5, etc.

Each disease status for individuals was based on the exposure level (present / absent), the background disease risk, and the relative risk. For example, if the background disease risk was 0.1, the relative risk of Disease 3 was 2.0, so the risk was the product, 0.2. Individuals were assigned Disease 3 status randomly, with binomial probability of 0.2. When the background prevalence and the RR were high, the product could produce a probability greater than 1, so we ‘capped’ probabilities at 0.9. As with prevalence, the actual RRs differed from the target values.

Three additional populations were created with different prevalences but no variation in the parameters across or within the towns. These provided a ‘control’ for our procedures.

### Applying the sampling methods

The methods all used a cluster sampling design. Apart from SA, the PSUs were towns. Thirty PSUs were selected using Probability Proportional to Size (PPS). In practice, this is Probability Proportional to Estimated Size (PPES) since the PSU sizes are not known exactly. We used two ways to identify PSUs for the simulations. The first (‘same PSUs’) identified 30 PSUs which were used to obtain all 1,000 sets of simulated samples. The second approach (‘resampling’) took a fresh sample of PSUs each time a new set of samples was taken. A set consisted of the three samples sizes (210, 450, and 900) x five sampling methods, i.e., 15 samples. One thousand sets of samples were taken.

Households were selected in each PSU until the specified sample size of individuals was reached. Sometimes the PPS selection method chose a town more than once. If the town was chosen k times, then k samples were taken from the town.

The sampling methods within the PSUs were:

#### Simple random sampling – ‘Random’

Simple random sampling (SRS) selected households with equal probability within PSUs. While logistically impractical in real-life populations, SRS was our standard for comparisons of the methods.

#### The original EPI method – ‘EPI’

We followed the original Extended Program on Immunization (EPI) random-walk approach [World Health Organization, 2005] described above. We used the centre of the town in place of a landmark. In practice buildings occupy an area in two dimensions, whereas we placed each building at a point. So instead of drawing a line from the centre of the town to the edge, we drew a narrow strip, symmetrical about the random direction, and identified buildings in that strip. We randomly chose one as the starting household and identified nearest neighbors until the required sample size was achieved (See Appendix figure).

#### Selecting parts of the sample from each quadrant – ‘Quad’

We divided each selected town into four quadrants and applied the original EPI method to each of them, replacing the central landmarks with the centres of the quadrant. Bennett et al. [1994] took a quarter of the sample from each quadrant. Our sample sizes per town were not divisible by four, so we ensured the sample size per quadrant was as even as possible, randomly determining which areas would have an extra ‘participant’.

#### Square grid – ‘Square’

We constructed a 64 × 64 grid of squares over each town. We randomly sampled squares, then one household within each square, and continued until the required sample size was reached.

#### Small areas as PSUs – ‘SA’

We constructed SAs by dividing towns into rectangular areas with between 50 and 100 households. SAs were chosen randomly from the whole population using probability proportional to size and households were randomly selected from each of the selected EAs until the target sample size was attained.

#### Sample size per PSU

Within each town (or SA), for each sampling method we used three sample sizes: 7, 15, and 30 children. The samples were chosen independently, and yielded overall sample sizes of 210, 450, and 900. For each sample size, we conducted 1000 simulations of the sampling.

### Analysis

#### Calculating probabilities of selection

The original EPI methodology treats samples within towns as simple random. Under this assumption, since towns are selected with probability proportional to size, these samples are self-weighting, i.e., the probability of selecting any individual is constant. We assumed this property also applied for Quad and SA. For the Square method, we estimated the overall probability of selecting an individual in the sample by multiplying together the probabilities of selecting the town, selecting the squares within the town (accounting for empty squares), and the household within the square (accounting for households with no children). The sampling weight was the inverse of this overall probability.

#### Calculating Prevalences and Relative Risks

For each sample size (210, 450, or 900) we computed the four prevalences of disease and the RRs, applying sampling weights when appropriate, for each of the 1000 simulations. Since the true prevalences and RRs were known, we computed the error of each sample (sample estimate minus true value) and took the mean of those values to estimate the bias.

We computed the variance of the estimates across the 1000 simulations. We used the bias and variance to compute the Mean Squared Error (MSE), where

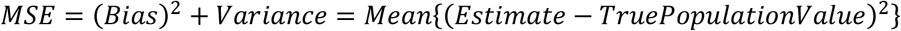

The Root Mean Squared Error (RMSE), the square root of the MSE, was our measure for comparing the sampling methods.

#### Overall comparisons of the sampling methods

We compared the sampling methods in two ways: firstly, for each population (and sample size) we ranked the RMSEs for the four methods. Lower RMSEs had lower ranks. We calculated the mean rank for each sampling method across the 50 populations.

Secondly, for each population (and sample size) we took the ratio of the RMSE for the sampling method to the RMSE for simple random sampling, our gold standard. We calculated the mean of these ratios for the 50 populations and compared the means between the sampling methods.

#### Impact of the population parameters

We also wanted to learn how the RMSE varied with different values of the parameters used to construct the populations. We created graphs showing the RMSEs for the different methods in relation to the parameter values. We smoothed the plots using generalized additive models.

### Computing

The creation of the populations and simulations of sampling were conducted on a modern high-performance cluster: we used SHARCNET, a computational resource supported by a consortium of Ontario universities [Anon, n.d.]. The two runs used for our final data took approximately 380 processor hours.

## Results

Overall analyses of RMSE Ratios and their ranks

### Mean ranks

Tables 1 and 2 show the mean ranks for when the Relative Risk was 1.0 and 3.0, respectively, and the same towns were ‘reused’ for each of the 1000 simulations. The results for other situations are similar and shown in the Supplementary information.

**Table 1.**
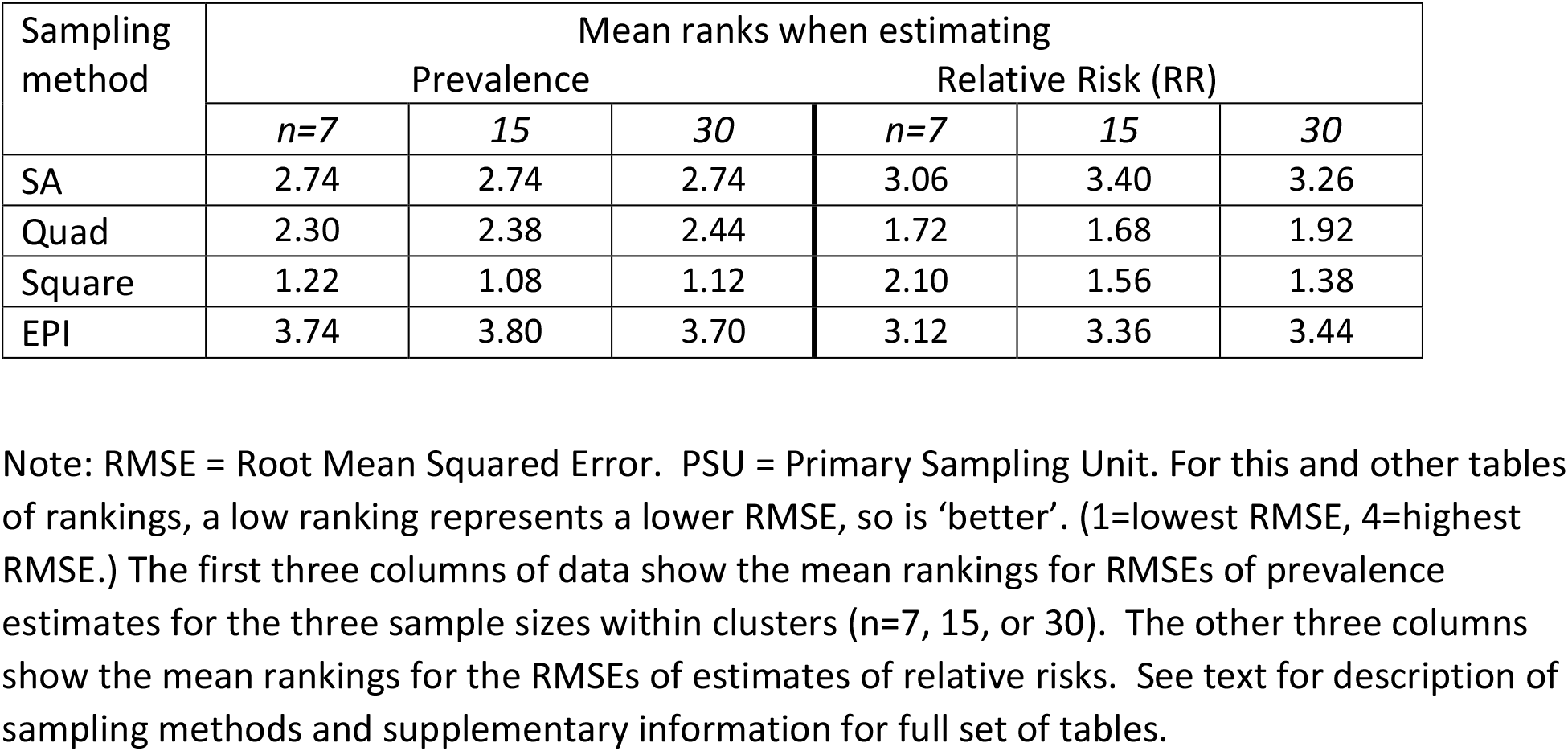
Mean ranks of RMSEs for relative risk = 1.0 and same PSUs are sampled

**Table 2.**
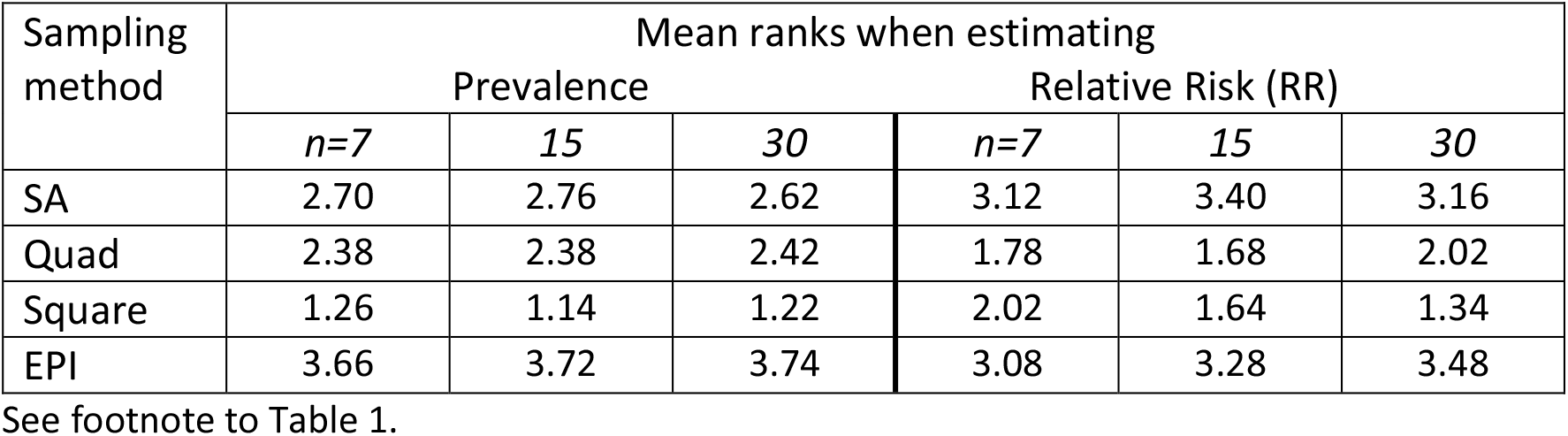
Mean ranks of RMSEs for relative risk = 3.0 and same PSUs are sampled

For estimates of prevalence, the Square method was the best, with mean rankings lower than (i.e., better than) those of other methods. Indeed, it ranked the best for at least 40 of the 50 populations regardless of the sample size or the sampling of towns. SA and Quad were similar. The EPI method was generally worse. Overall, the mean rankings did not change much with sample size.

For estimates of Relative Risk, the picture is a little different. For the sample sizes of 7 per PSU, the Quad method had the lowest mean ranks. For 15 per PSUr, the mean ranks for Quad and Square methods were very similar. For the largest sample size (30 per PSU) the Square technique was the best.

### Means of ratios of RMSEs

The means of the ratios of RMSEs (to the RMSEs for simple random sampling) are shown in Tables 3 and 4 for Relative Risks of 1.0 and 3.0 when the same towns were used for each of the 1,000 simulations. Once again, results for other cases are similar and are included in the Supplementary information. We also examined the results graphically (Figure 1). Part (a) shows RMSE ratios when estimating prevalence for RR=1.0 and the same towns were used for the simulations and is typical of the graphs for the other conditions. Part (b) shows the results when estimating RR under the same conditions. Other graphs (in the Supplementary information) show mostly similar patterns reflecting the results seen in Tables 3 and 4.

**Table 3.**
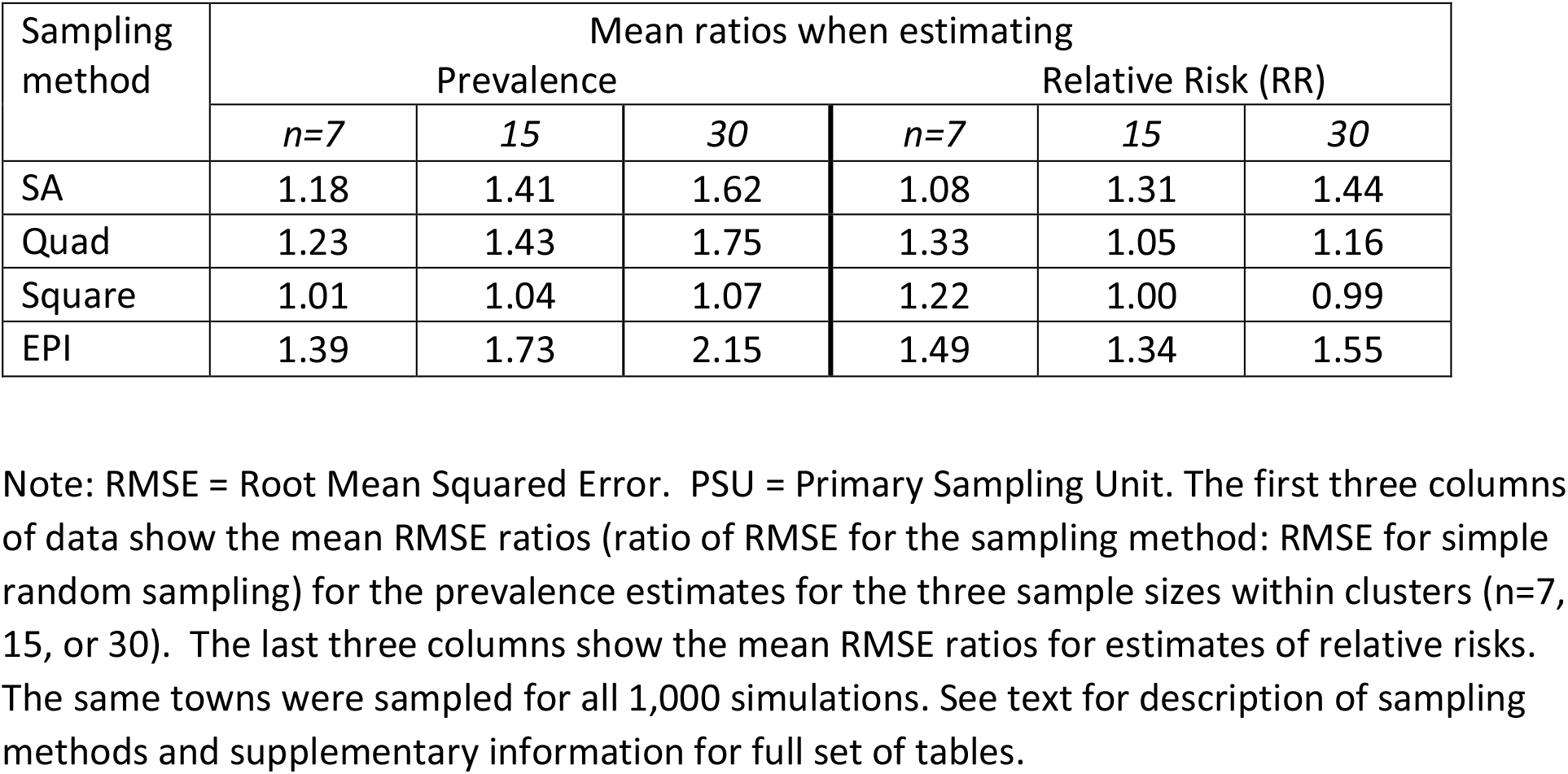
Mean ratios of RMSEs for relative risk = 1.0 and same PSUs are sampled

**Table 4.**
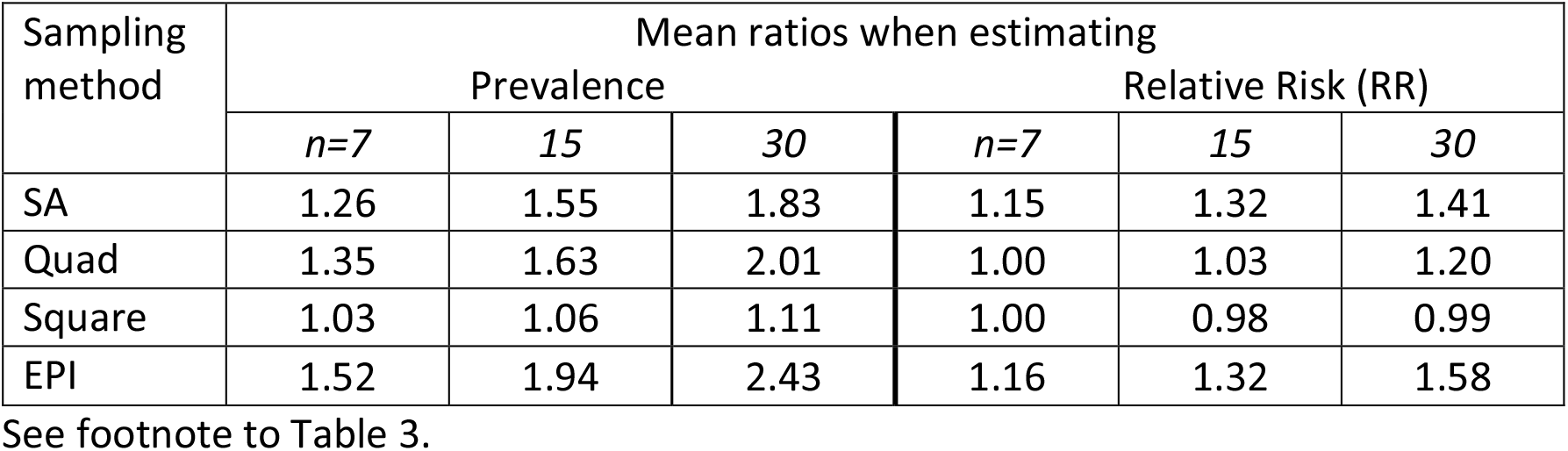
Mean ratios of RMSEs for relative risk = 3.0 and same PSUs are sampled

**Figure 1.**
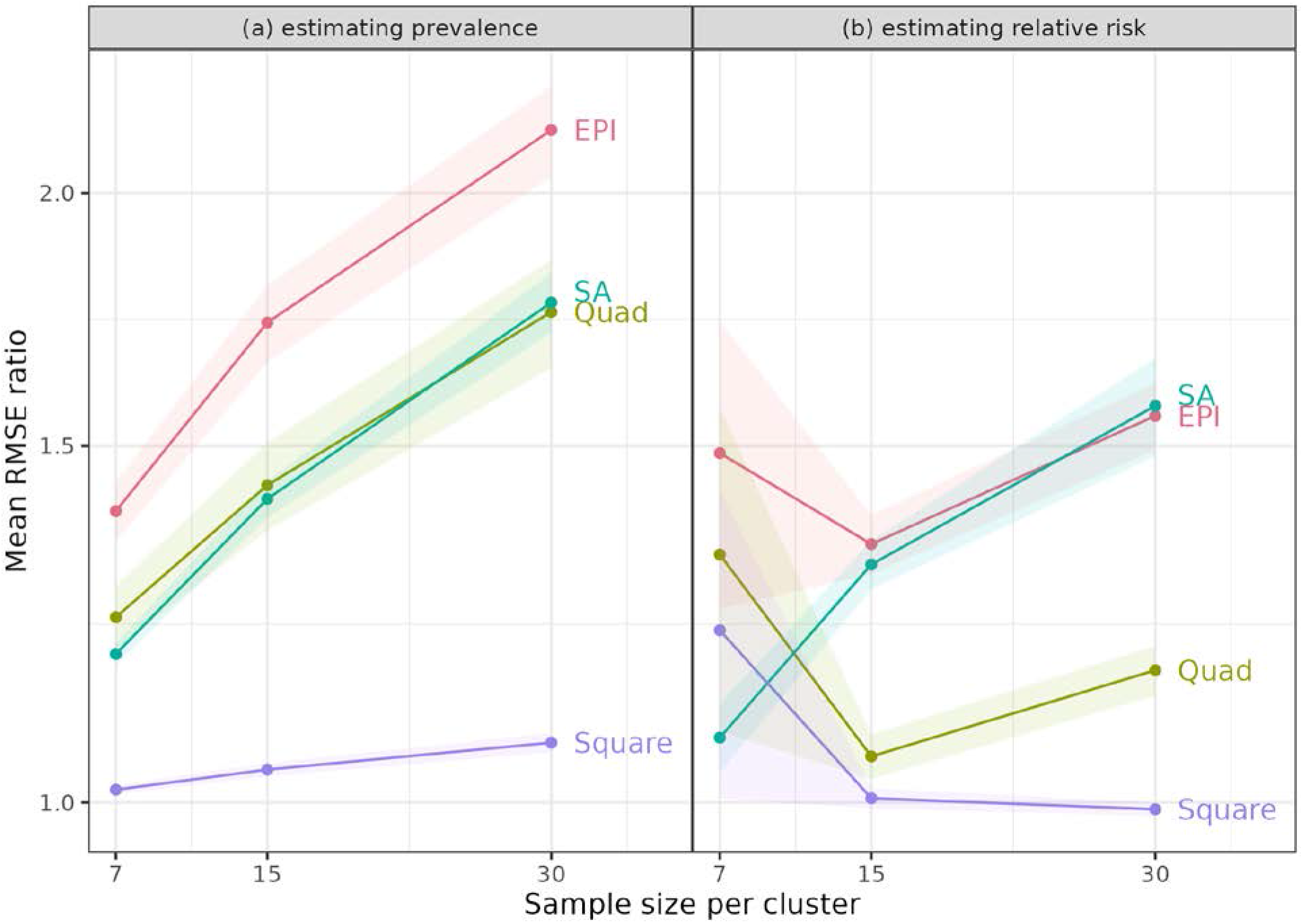
Mean of ratios of RMSE for sampling method to RMSE for simple random sampling. Figure shows the mean ratios when estimating (a) Prevalence and (b) Relative Risk (RR), using the same sample of towns (clusters for the SA method) for each population, and RR=1.0. PSU = Primary Sampling Unit.

For estimating prevalence, the Square method was always best – it had the lowest mean ratios, which were close to 1 for all sample sizes, indicating that the RMSEs were similar to those from simple random sampling (SRS). Notably, the other methods had mean ratios that increased with sample size per PSU. With SRS, statistical theory predicts that an increase in sample size from 7 to 30 per PSU will reduce the variance of estimates by a factor of just under a quarter (7/30). The increase in the ratios indicated that these methods benefited less from larger sample sizes. This disadvantage likely reflects some intracluster correlation due to the homogeneity of people in neighbourhoods. This result was not surprising, since these methods sample close neighbours within clusters.

One might have expected the Quad approach to be relatively free of this property, since it samples from different areas of the PSUs, but it also showed an increase in the mean ratio with larger sample size. Since SA takes random samples, it might have avoided the problem, but it did not.

For estimates of Relative Risk, the Square method performed very well; the mean RMSE ratios were mostly close to 1.0, for all three sample sizes. The Quad procedure was sometimes - but not always – comparable in having low mean ratios.

### Impact of parameter values

Given the results above, we did not expect that examining the relationship between the RMSEs and parameter values (which characterized the populations) would identify circumstances when a method other than Square would be preferable. Still, for completeness, we looked at the relationships. We examined graphs of the mean RMSE ratios as a function of parameter values (Figure 2).

**Figure 2.**
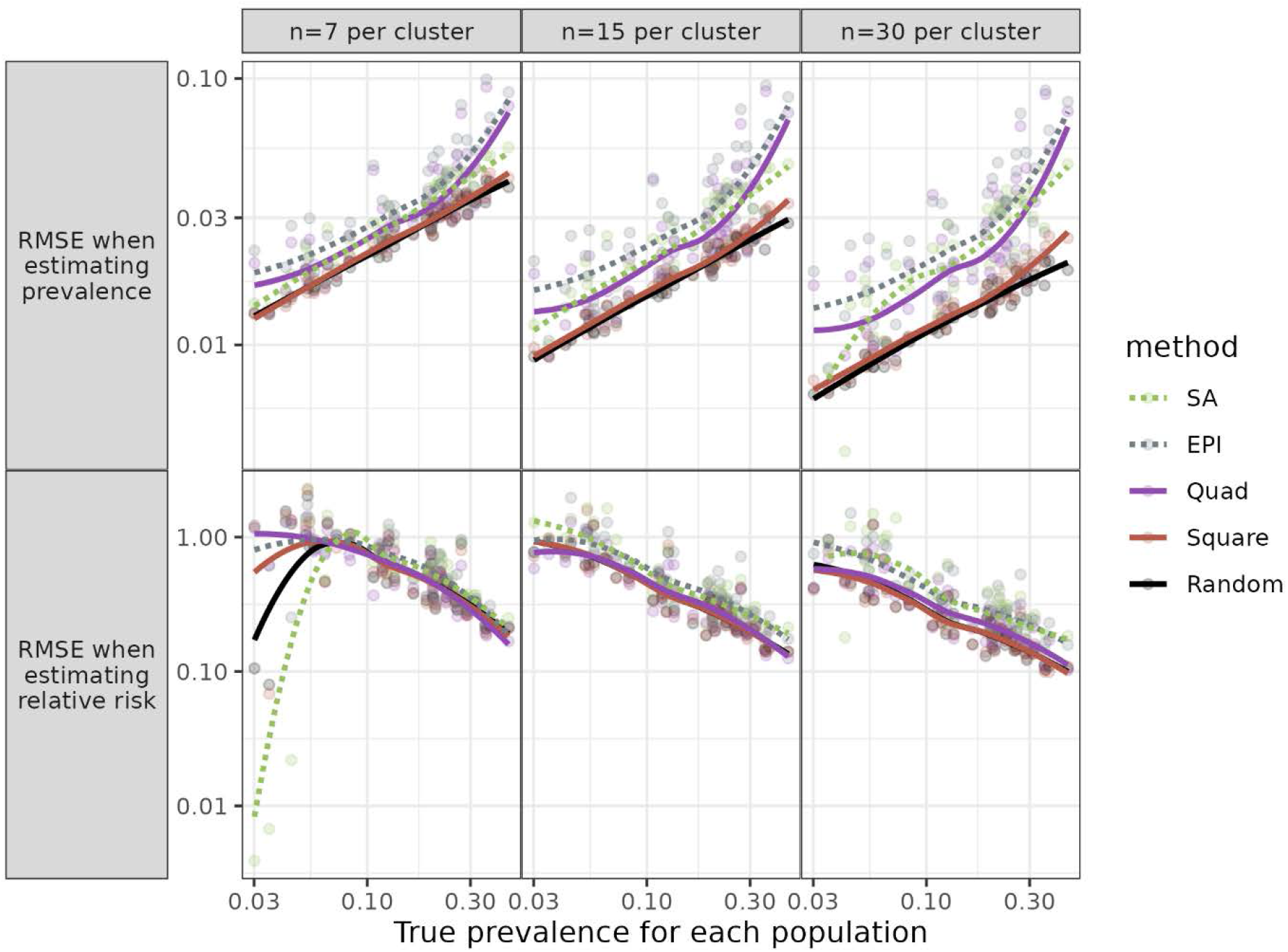
Root Mean Square Error (RMSE) for each population against prevalence. Figure shows RMSE for three sample sizes, when the same towns were sampled for each simulation and relative risk (RR) = 1.0. PSU = Primary Sampling Unit.

Individual parameters had little or no impact on the relative performance of different methods when estimating prevalence. This was mostly the case for estimates of RR. Especially for the larger sample sizes (n=15 or 30 per PSU) the relative values for the different methods were mostly independent of parameter values.

Further details of the Results are in the Supplementary information.

### Non-varying populations

For the three populations for which all individuals had the same probability of disease, all methods were similar in their RMSEs (data in Supplementary information).

## Discussion

### Summary of main results

Our simulations found that the Square method was nearly always the best, as measured by lower RMSEs. Under some circumstances, the Quad approach, which samples from four areas of each town, performed quite well, better than the EPI method, but not as well as the Square technique. SA was mostly an improvement over EPI, especially when estimating prevalence. The other criterion for comparison, the ranks of RMSE ratios, suggested that the Square method was almost universally better.

The examination of RMSEs in relation to population parameters revealed that there were no particular parameters (i.e., no population types) for which the relative ranking of the methods varied, at least for the larger sample sizes. For the three non-varying populations, as expected, there were minimal differences between methods.

### Commentary

Several procedures have been proposed to overcome the known limitations of the original EPI. These new procedures did improve on EPI but had their own limitations. Thus, some authors [e.g., 15] have proposed segmenting towns into smaller units, whose populations can be enumerated to allow simple random sampling. Our results for the SA approach, though, suggest that the homogeneity within small segments produces sufficiently large design effects that increasing sample size within the segments does not improve precision as much as expected. Moreover, it requires some prior identification of the SAs, beyond data on town population sizes alone.

Designers of those surveys were well aware of the impact of clustering. The Reference manual for the new WHO EPI method (what we labelled SA) includes a table of the design effects (DEFF) with different values of the Intracluster Correlation Coefficient (ICC) [8:127]. It states that a conservative estimate of the ICC for routine immunization surveys is 1/3, or 0.333. With seven respondents per PSU (cluster) the DEFF is 3.0, so that three times as many SAs must be sampled to achieve the same precision as a random sample. This would add considerably to the time and/or cost to do the study.

The Square approach, which does not have this limitation, could be adapted in the absence of information on the target population, for example, when an informal refugee camp is formed. Drones or other technology could ensure the aerial images used are up-to-date. This approach would be even more feasible if newer software can recognize buildings or tents on the ground, so the step of identifying structures could be automated.

One possible disadvantage of the Square method is that for larger towns, there may be a good deal of travel (hence increased cost) required to reach all the sampled households, while the other methods restrict samples to a small geographic area. Still, at times this may be an advantage if there are concerns about the security of interviewers: with the Square method, interviewers can enter and leave areas quickly, rather than spending time finding and interviewing several households in a small neighbourhood.

### Strengths and limitations of our work

Our study has several strengths. We attempted to create realistic populations, whose characteristics varied between and within towns. We included multiple populations, which simulations using real data cannot. Our full analysis included many sampling methods, including some variations on EPI that have been proposed but to our knowledge have not been used in practice. For the SA and Square techniques, sampling probabilities can be properly estimated, unlike the EPI method (and its variants).

Of course, our study also has limitations. The populations are simulated, not real. Small neighbourhoods in our simulations may be more homogeneous than in real life; still, similarity of nearby households is broadly realistic. Our simulated samples were ideal and ignored the logistical difficulties experienced by real surveys. For example, population numbers are inexact so PPS sampling is subject to error; interviewer teams make decisions that may not strictly follow protocols; and people in households may be out when interviewers call or may refuse to participate. (As noted earlier, Cutts and colleagues [12] provided a fuller discussion.) Still, we expect these problems would apply similarly - and lead to similar degrees of inaccuracy - for different sampling methods. In practice, the Square approach relies on some technical ability to deal with images and to identify GPS locations of buildings. It also requires identifying buildings from aerial images, which can lead to errors due to, e.g., tree coverage.

The time required to complete the survey may influence the choice of sampling method. EPI and its variants can be completed quickly, while the WHO manual for the updated EPI methodology (i.e., SA) projects an overall 12-month timetable [8:23]. The Square method requires obtaining the relevant images and identifying buildings from them, which should be possible to do quite quickly: a sample of the grid squares can be chosen and surveyors need only identify buildings in those squares.

### Contribution of our study

Our work adds to the literature in several ways. To our knowledge, it is the first simulation study to explore the properties of small area (SA) sampling and ‘Square’ sampling. Given modern computing power, we were able to simulate large populations across hundreds of towns. We varied parameters across these towns to create more realistic populations and examined the impact of these parameters. We compared multiple sampling methods. We know of no other study that evaluates how different sampling methods affect estimates of relative risk. Finally, we included the previously-untested Square method, which has proved to be statistically superior to other sampling approaches that are used in several major official surveys.

### Conclusion

The simulations show that the Square method is almost always the best from a statistical perspective, especially when estimating prevalence or for larger sample sizes. Quad and SA improve on the original EPI (EPI), but not enough to be statistically preferable to the Square method, which is relatively easy to apply.

## Supporting information

Details of main results

Details of main results including some sampling methods not included in the paper

Various extra information including more methodological details and results

## Data Availability

Since ours was a simulation study in which we created the populations, we do not have data per se. The software for creating the populations and running the sampling methods was written primarily by one of us (PDE).
Project home page: https://github.com/patrickdemond/sampsim/
Archived version: The source code can be downloaded at http://doi.org/10.5281/zenodo.4731010
Operating system(s): Platform independent
Programming language: R, C++
Other requirements: cmake, and LibArchive, JSON Cpp, and C++11 compliant compiler must be linked for the project to compile
License: GNU General Public License 3.0. https://choosealicense.com/licenses/gpl-3.0/
Any restrictions to use by non-academics: No
The supplementary information, uploaded with the manuscript, contains excel files of the output from the program.

## Author contributions

**Conceptualization:** Harry S. Shannon, Román Viveros-Aguilera

**Formal analysis:** Patrick D. Emond, Harry S. Shannon

**Funding acquisition:** Harry S. Shannon, Román Viveros-Aguilera, Benjamin M. Bolker

**Investigation:**

**Methodology**: Harry S. Shannon, Román Viveros-Aguilera, Benjamin M. Bolker, Patrick D. Emond

**Project administration**: Harry S. Shannon

**Software:** Patrick D. Emond, Benjamin M. Bolker

**Supervision:** Harry S. Shannon

**Visualization:** Benjamin M. Bolker

**Writing – original draft:** Harry S. Shannon

**Writing – review & editing:** Harry S. Shannon, Román Viveros-Aguilera, Benjamin M. Bolker, Patrick D. Emond

## Declarations

### Ethics approval

Since no humans or their data were used, ethical approval was not required.

### Author Contributions

HSS and RV-A conceived the study, and HSS, RV-A, and BB developed the protocol. PDE programmed and undertook the simulations, with input from BB, HSS, and RV-A. HSS wrote the initial draft of the manuscript. All authors contributed to revisions and read and approved the final manuscript.

### Data availability

Since ours was a simulation study in which we created the populations, we do not have data per se. The software for creating the populations and running the sampling methods was written primarily by one of us (PDE).

- Project home page: https://github.com/patrickdemond/sampsim/
- Archived version: The source code can be downloaded at http://doi.org/10.5281/ze-nodo.4731010
- Operating system(s): Platform independent
- Programming language: R, C++
- Other requirements: cmake, and LibArchive, JSON Cpp, and C++11 compliant compiler must be linked for the project to compile
- License: GNU General Public License 3.0. https://choosealicense.com/licenses/gpl-3.0/
- Any restrictions to use by non-academics: No

### Supplementary information

The program output is included in the supplementary information as two Excel files. We also include a file that provides more methodological details and extra tables, as well as a section that explains how to interpret the Excel files.

### Funding

The study was funded by the Canadian Institutes of Health Research, Funding Reference Number: 123432.

## Acknowledgements

Not applicable

## Conflict of Interest

None declared

## Appendix: Diagrams showing sampling methods

Each diagram shows a town. For simplicity of presentation, we sample three households per town (or per Small Area).

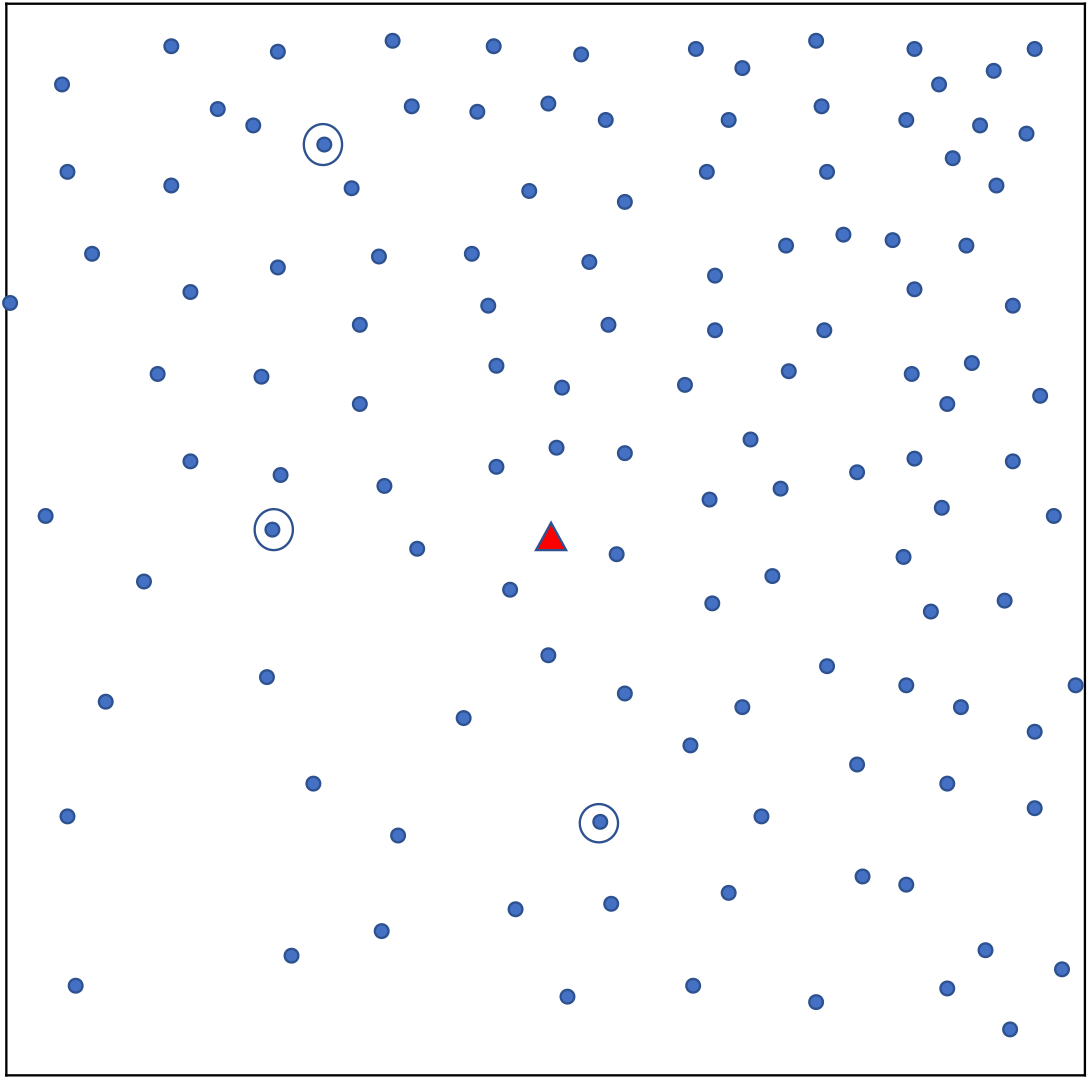

Simple random sampling. Each dot shows a household. The triangle represents a central landmark. Three households (circled) are randomly chosen.

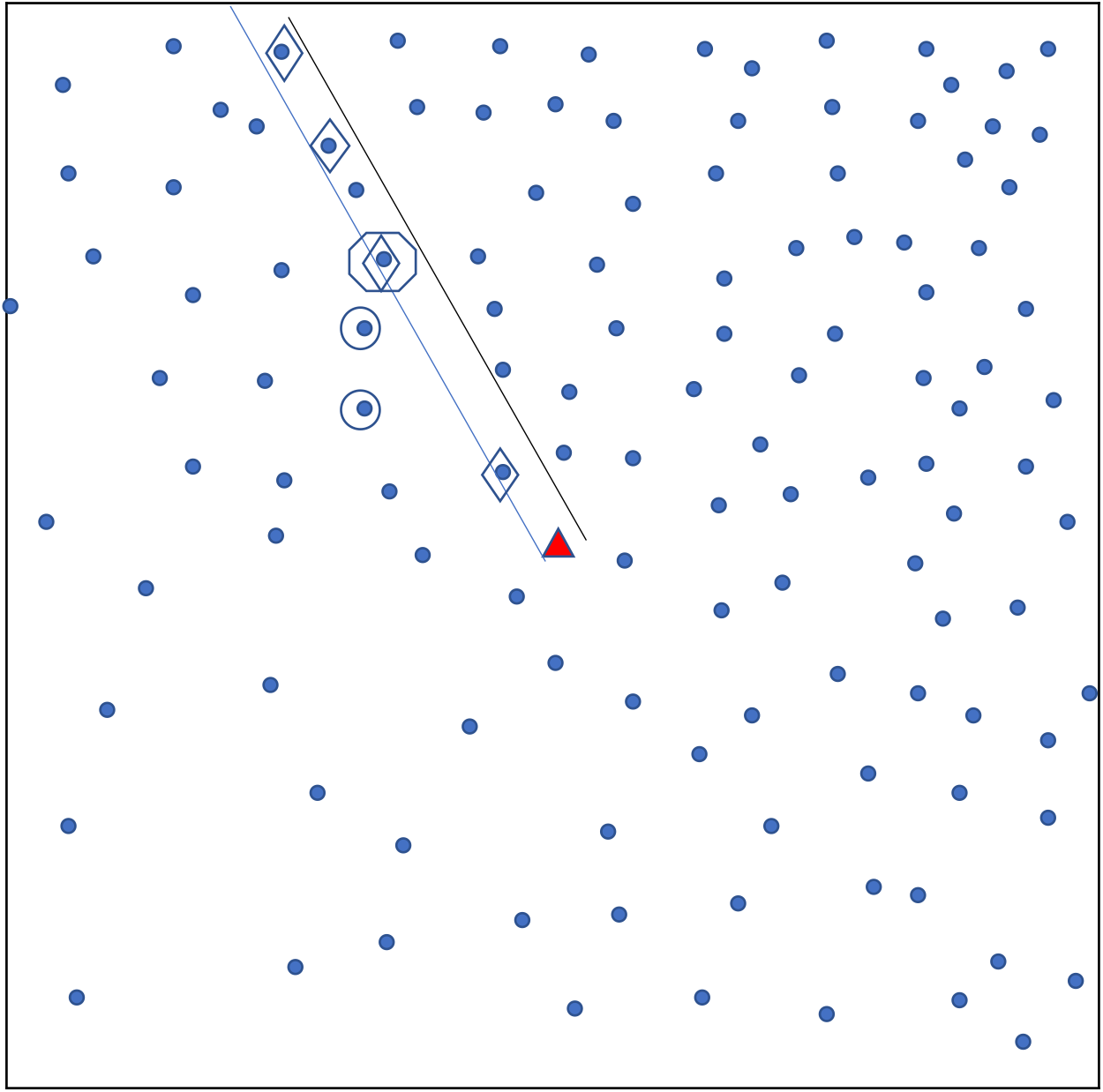

Sampling using original EPI method. Each dot shows a household. A central landmark is identified (triangle). A random direction is chosen (parallel lines) from the landmark and households in that direction are identified (diamonds). From these, the ‘starting’ household’ is randomly chosen (octagon) and nearest neighbours are also selected for the sample (circles).

The ‘Quad’ sampling method divides a town into four quadrants and applies this sampling approach in each quadrant.

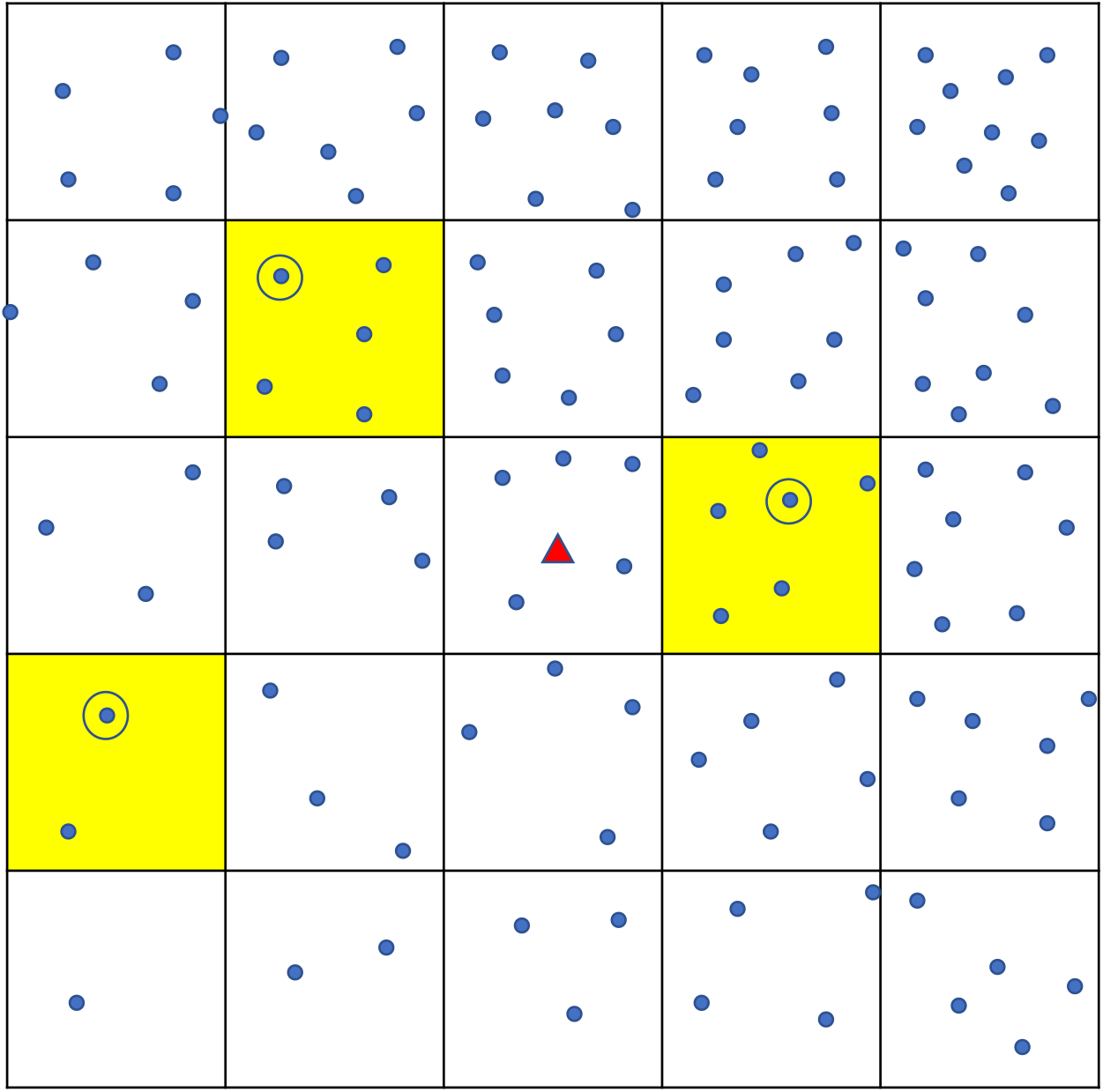

Sampling using ‘Square’ method. Each dot shows a household. The town is divided into a grid of smaller squares. Yellow shading shows the three squaresthat are randomly chosen, and one household (circled) is chosen from each.

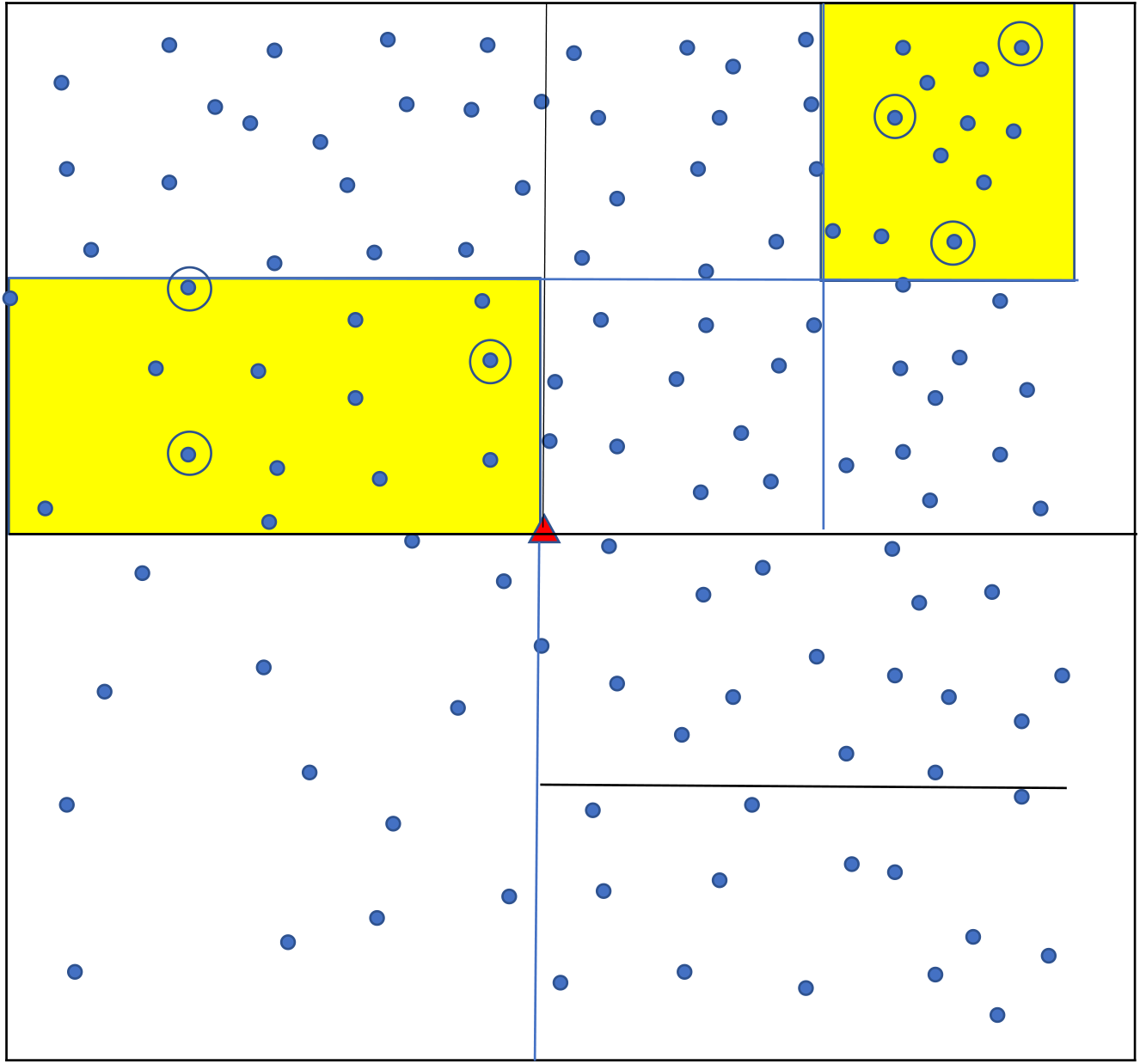

Small area (SA) sampling. The town is divided into successively smaller areas until each contains a number of households in the pre-specified range. Several small areas (yellow shading) are randomly chosen. Three households are randomly sampled from each selected small area.

## Notes

### Competing Interest Statement

The authors have declared no competing interest.

