## Supplementary material for "A Simulation Study of Sampling in Difficult Settings: Statistical Superiority of Little-Used method": Various extra information including more methodological details and results

**Contents of supplementary information**

This file contains:

- An introduction
- A list of parameters used in creation of populations and disease determination
- The disease determination algorithm
- Further details on the sampling methods
- Supplementary tables – complete set of tables
- A description of how to interpret the Excel files of results

***Introduction***

The supplementary material includes more details on the creation of the populations, the sampling methods, and the results of the methods we did not present in the main manuscript. Apart from this file, there are two Excel files that contain the detailed results for each population we created and a zipped file with many figures.

The first Excel file (Results4x50.xlsx) is the one we used for the tables in the manuscript. The second (Results8x50.xlsx) adds other sampling methods we investigated. As we noted in the main document, for clarity we excluded several variants of the original EPI method for clarity, and only included the variant that performed best. This file includes a guide to understanding the Excel files.

***Parameters used in creation of populations and disease determination***

| **Label** | **Variable** | **Values used in this study** |
|---|---|---|
| seed | Seed used by random generator | |
| target_prevalence | Target disease prevalence | (0.1,0.5] |
| populations | Number of populations to generate | 50 |
| towns | Number of towns to generate | 300 |
| town_size_min | Minimum population of a town | 400 |
| town_size_max | Maximum population of a town | 300,000 |
| town_size_shape | Shape parameter used by town size Pareto distribution | 0.785 |
| tile_x | Number of tiles in the horizontal direction | 10 |
| tile_y | Number of tiles in the vertical direction | 10 |
| tile_width | Width of a tile in kilometers | 1 |
| popdens_mx | Population density trend's X coefficient | (0,1.0] |
| popdens_my | Population density trend's Y coefficient (must be [-1,1]) | (0,1.0] |
| mean_household_pop | Mean number of individuals per household | (2,5] |
| disease_pockets | Number of disease pockets to generate per town | [0,10], Integer values only |
| pocket_kernel_type | Type of kernel to use for disease pockets | Exponential; Inverse square; Gaussian |
| pocket_scaling | The scaling factor to use for disease pocket | (0.5,2] |
| mean_income_b00 | Mean income trend's base value | See array in Excel file 'Parameters' sheet |

| | | |
|---|---|---|
| mean_income_b01 | Mean income trend's X coefficient base value | See array in Excel file ‘Parameters’ sheet |
| mean_income_b10 | Mean income trend's Y coefficient base value | See array in Excel file ‘Parameters’ sheet |
| sd_income_b00 | SD of values of income | 0.25 |
| mean_disease_b00 | Mean disease trend's base value | See array in Excel file ‘Parameters’ sheet |
| mean_disease_b01 | Mean disease trend's X coefficient base value | See array in Excel file ‘Parameters’ sheet |
| mean_disease_b10 | Mean disease trend's Y coefficient base value | See array in Excel file ‘Parameters’ sheet |
| mean_exposure_b00 | Mean exposure trend's base value | See array in Excel file ‘Parameters’ sheet |
| mean_exposure_b01 | Mean exposure trend's X coefficient base value | See array in Excel file ‘Parameters’ sheet |
| mean_exposure_b10 | Mean exposure trend's Y coefficient base value | See array in Excel file ‘Parameters’ sheet |
| dweight_income | Disease weight for household income | (0,1] |
| dweight_risk | Disease weight for household risk | (0,1] |
| dweight_age | Disease weight for household age | (0,1] |
| dweight_pocket | Disease weight for pocketing | 1 |

Notes: Coefficients for Income, Disease, and Exposure were for use in linear function based on position of household in town based on X and Y coordinates. Disease weights were applied in the Disease Determination Algorithm. For values shown as a range, the Latin Hypercube selected the 50 values at equal intervals between the lowest and highest values of the range. See ‘Creating the virtual populations’ in the text for more detail.

### Disease determination algorithm

*Written by Patrick D. Emond*

The probability of disease for each individual is determined by the following equation:

$$P(i) = \frac{1}{1+e^{-\eta(i)}}$$

where $i$ is a particular individual and η is the sum of all weighted factors:

$$\eta(i) = \sum_f W_f Z_f(i)$$

where $f$ represents each disease factor, $W_f$ is the weight for the disease factor $f$ and $Z_f(i)$ is the normalized value for the factor $f$ for the individual $i$ :

$$Z_f(i) = \frac{X_f(i) - \mu_f}{\sigma_f}$$

where $X_f(i)$ is the individual's value for factor $f$, and $\mu_f$ and $\sigma_f$ are the mean and standard deviation, respectively, of all values for factor $f$. Combining these into one equation, this resolves to

$$P(i) = \left\{1 + exp\left[-\sum_f W_f \frac{X_f(i) - \mu_f}{\sigma_f}\right]\right\}^{-1}$$

The six factors are population, income, disease risk, age, sex, and pocketing:

$$X_{population} \equiv N_{household}$$

$$X_{income} \equiv -ln\mathcal{N}\{F_\mu()|_\square, F_\sigma()|_\square\}$$

$X_{age} \equiv 0$, if child
1, if adult

$X_{sex} \equiv 0$, if female
1, if male

$X_{pocket} \equiv \sum_{pocket} e^{-d_p}$ , if exponential
$\equiv \sum_{pocket} {d_p}^{-2}$ , if inverse square
$\equiv \sum_{pocket} e^{-{d_p}^2}$ , if Gaussian

where $ln\mathcal{N}(\mu, \sigma)$ and $\mathcal{N}(\mu, \sigma)$ are the log-normal and normal distributions for mean μ and standard deviation σ, $F()$ is a spatial function of the form $F() = b_{00} + b_{10}x + b_{01}y$ , $F()|_{\boxdot}$ is the function $F()$ evaluated at the centroid of a tile and $d_f$ is the distance from a household to the pocket *p* divided by a programmable scaling constant.

By default this algorithm resulted in the mean disease prevalence always being near 0.5. However, it was possible to change the resulting mean to a target mean $\tau$ by shifting the function for P such that it crossed the Y-axis at $\tau$. First we had to solve *P* for $\eta$:

$$\eta = -ln\left(\frac{1}{p} - 1\right)$$

then add the value of $\eta$ for $P = \tau$ from the function for $\eta(i)$:

$$\eta(i) = \sum_f W_f Z_f(i) - ln\left(\frac{1}{\tau} - 1\right)$$

Finally, the above adjustment to $\eta$ will result in a mean prevalence that follows an arc-sine curve. To get the desired mean prevalence the value for $\tau$ must be adjusted by an ad hoc function:

$$\tau_0 = \frac{1}{2}\left\{sin\left[\frac{\pi}{2}(2\tau - 1)\right] + 1\right\}$$

***Further details on the sampling methods and their operationalization***

The methods all used a cluster sampling design. In all methods except one (labelled 'SA'), the primary sampling units (PSUs) were towns. The sampling methods within the PSUs were:

*Simple random sampling – 'Random'*

Simple random sampling (SRS) selects households with equal probability within PSUs. While logistically impractical in real-life populations, SRS was our standard for comparisons of the methods.

*The original EPI method – 'EPI'*

The original Extended Program on Immunization (EPI) approach identified a landmark in the centre of a town, and chose a random direction from that landmark. Interviewers walked along that line to the edge of the town, identifying buildings on the line. One building was randomly chosen as the starting point. The household in that building (if any) was asked to participate in the survey. The next household chosen was the next closest household residence. This 'nearest neighbour' identification continued until the required sample size was reached. EPI aimed to estimate the proportion of young children who had been immunized against some disease(s), so limited interviews to households with eligible children.

In practice buildings occupy an area in two dimensions, whereas we placed each building at a point. So instead of drawing a line from the centre of the town to the edge, we drew a strip 0.1 units wide (one hundredth of the length of each side of the town), symmetrical about the random direction, and identified buildings in that strip.

*The EPI method, choosing every 3rd or 5th household – 'EPI3', 'EPI5'*

By choosing every 3rd or 5th household, it is expected that the sample will come from a wider geographical area, and thereby be less susceptible to biases [Bennett et al]. We operationalized this approach by using the EPI method of successively identifying the nearest neighbour, but only sampling every 3rd or 5th household.

(For completeness, when sampling via EPI, EPI3, and EPI5, we determined an arc of 6°, symmetrical about the random direction. One building in the strip or arc was randomly selected as the starting point. We found almost no difference between the results for the two approaches, so we only consider the strip selections.)

*Half sample from centre, half from periphery – 'Peri'*

This method [Bennett] also aims to obtain the sample from a wider geographic area than EPI. A random direction is chosen from the centre of a town, and the first household in that direction is sampled. The nearest neighbour method then obtains half the target sample for that town. A new random direction is found and the last household along that line to the edge of the town is sampled. The remainder of the sample is found using the nearest neighbour approach. We used strips rather than lines as described above to identify the two 'starting' households.

*Selecting parts of the sample from each quadrant – 'Quad'*

A further method designed to provide geographic dispersion of the sample divides the town into four quadrants, and applies the OldEPI method to each of them, replacing the central landmarks with the centres of the quadrants [Bennett]. A quarter of the sample is taken from each quadrant.

The Peri and Quad simulations did not yield equal numbers in the different areas when the target sample size in any town was not divisible by two (Peri) or four (Quad). We ensured the split was as even as possible, randomly determining which areas would have an extra ‘participant’.

*Using a grid to identify the initial household - ‘Grid’*

One approach that avoids the use of a central landmark was to use a rectangular grid ‘superimposed onto a scanned streetmap of the quartier,’ randomly select an intersection of the grid lines, and identify the ‘closest compound to the right’ as the starting point for the EPI method [Grais].

*Circles about random points - ‘Circle’*

Random GPS points are chosen in each town, a circle of fixed radius is drawn on an aerial image around each point, the buildings within the circle are identified from the image, one building is randomly chosen, and a household in the building is interviewed. This continues until the target sample size is reached [Kolbe]. We applied this technique, using circles of radius 0.1 units.

*Square grid – ‘Square’*

The potential for overlapping circles in the above procedure complicates the estimation of probabilities of selection. Instead, a grid of squares can be imposed on the image of the town to avoid overlap. (In practice, adjustment to town boundaries is necessary.) Grid squares can be randomly chosen, and a household randomly selected from those squares [Shannon]. In this study, we used a 64 x 64 grid of squares over each town. In practice, buildings can overlap the edges of the squares. Since we defined buildings as points, we did not have this problem, although buildings could land on an edge dividing squares. We specified that a building on the south or west edge of a square was deemed to be in that square.

*The small area method – 'SA'*

Several surveys use a different method of sampling than EPI [MICS, DHS]. In brief, the approach is as follows: The population under study is divided into Enumeration Areas (or equivalent), which are of fairly similar size. Large EAs are segmented into smaller areas, while small EAs are combined with contiguous areas to ensure that the EAs differ in size by no more than twofold.

EAs are then selected using PPES. Once the sample of EAs has been chosen, the households in each are enumerated. A random sample within each EA is chosen, up to the required sample size.

The method differs from the other approaches, which all use towns as the clusters. By enumerating all households in the EAs, the probability of selection can be estimated, unlike in EPI and its variants. In our virtual populations, we did not create EAs. Instead, we constructed EAs by dividing towns into rectangular areas with between 50 and 100 households.

Other researchers have proposed alternatives to EAs, since the most recent censuses in lower-income countries may be very out-of-date. Several 'gridded' population datasets have been constructed. They use a statistical model with available spatial data to estimate populations in small grid cells. Thomson et al. describe 43 surveys that have used gridded datasets. Since these studies identify PSUs analogous to EAs, these gridded population surveys can be considered equivalent to the SA methodology for our purposes.

Yet another approach is to ‘segment’ clusters [Brogan, Turner]. The segmentation process is done by teams in the field. The teams produce rough maps of the clusters and divide them into smaller areas (segments) of roughly equal size. One segment is randomly chosen, and a complete list of households obtained, from which the required sample is randomly chosen. However, this procedure requires relatively small clusters, whose size is known from a recent census. Thus, in Turner et al.’s example, the clusters were mostly 250-300 households [Turner]. The SA technique uses EAs which likely match the sort of clusters needed for this approach, so we have in effect included sampling using segments.

In summary, we included 10 sampling methods: Random, EPI, EPI3, EPI5, Peri, Quad, Grid, Circle, Square, and SA. For our main document (i.e., our final paper) we include only five: Random, EPI, Quad, Square, and SA. The results for the Square and Circle methods were also very similar; we present results only for the former.

**Supplementary tables**

Note: these tables include and supplement Tables 1-4 in the manuscript.

**Tables of results with four sampling methods**

**Tables of Mean Ranks**

Table S1. Mean ranks of RMSEs for relative risk = 1.0 and same PSUs are sampled

| Sampling method | Mean ranks when estimating Prevalence | | | Relative Risk (RR) | | |
|---|---|---|---|---|---|---|
| | *n=7* | *15* | *30* | *n=7* | *15* | *30* |
| SA | 2.74 | 2.74 | 2.74 | 3.06 | 3.40 | 3.26 |
| Quad | 2.30 | 2.38 | 2.44 | 1.72 | 1.68 | 1.92 |
| Square | 1.22 | 1.08 | 1.12 | 2.10 | 1.56 | 1.38 |
| EPI | 3.74 | 3.80 | 3.70 | 3.12 | 3.36 | 3.44 |

Note: RMSE = Root Mean Squared Error. PSU = Primary Sampling Unit. For this and other tables of rankings, a low ranking represents a lower RMSE, so is 'better'. (1=lowest RMSE, 4=highest RMSE.) The first three columns of data show the mean rankings for RMSEs of prevalence estimates for the three sample sizes within clusters (n=7, 15, or 30). The other three columns show the mean rankings for the RMSEs of estimates of relative risks. See text for description of sampling methods.

Table S2. Mean ranks of RMSEs for relative risk = 1.5 and same PSUs are sampled

| Sampling method | Mean ranks when estimating Prevalence | | | Relative Risk (RR) | | |
|---|---|---|---|---|---|---|
| | *n=7* | *15* | *30* | *n=7* | *15* | *30* |
| SA | 2.72 | 2.78 | 2.80 | 2.94 | 3.38 | 3.18 |
| Quad | 2.28 | 2.40 | 2.42 | 1.84 | 1.60 | 2.00 |
| Square | 1.24 | 1.04 | 1.10 | 2.04 | 1.64 | 1.38 |
| EPI | 3.76 | 3.78 | 3.68 | 3.18 | 3.38 | 3.44 |

See footnote to Table S1

Table S3. Mean ranks of RMSEs for relative risk = 2.0 and same PSUs are sampled

| Sampling method | Mean ranks when estimating Prevalence | | | Relative Risk (RR) | | |
|---|---|---|---|---|---|---|
| | *n=7* | *15* | *30* | *n=7* | *15* | *30* |
| SA | 2.74 | 2.80 | 2.80 | 3.10 | 3.24 | 3.22 |
| Quad | 2.38 | 2.36 | 2.36 | 1.76 | 1.80 | 1.90 |
| Square | 1.14 | 1.12 | 1.12 | 2.04 | 1.62 | 1.42 |
| EPI | 3.74 | 3.72 | 3.72 | 3.10 | 3.34 | 3.46 |

See footnote to Table S1

Table S4. Mean ranks of RMSEs for relative risk = 3.0 and same PSUs are sampled

| Sampling method | Mean ranks when estimating Prevalence | | | Relative Risk (RR) | | |
|---|---|---|---|---|---|---|
| | *n=7* | *15* | *30* | *n=7* | *15* | *30* |
| SA | 2.70 | 2.76 | 2.62 | 3.12 | 3.40 | 3.16 |
| Quad | 2.38 | 2.38 | 2.42 | 1.78 | 1.68 | 2.02 |
| Square | 1.26 | 1.14 | 1.22 | 2.02 | 1.64 | 1.34 |
| EPI | 3.66 | 3.72 | 3.74 | 3.08 | 3.28 | 3.48 |

See footnote to Table S1

Table S5. Mean ranks of RMSEs for relative risk = 1.0 and different PSUs are sampled for each simulation

| Sampling method | Mean ranks when estimating Prevalence | | | Relative Risk (RR) | | |
|---|---|---|---|---|---|---|
| | *n=7* | *15* | *30* | *n=7* | *15* | *30* |
| SA | 2.58 | 2.68 | 2.62 | 3.28 | 3.50 | 3.36 |
| Quad | 2.36 | 2.48 | 2.44 | 1.48 | 1.58 | 1.78 |
| Square | 1.18 | 1.02 | 1.12 | 2.14 | 1.76 | 1.46 |
| EPI | 3.88 | 3.82 | 3.82 | 3.10 | 3.16 | 3.40 |

See footnote to Table S1

Table S6. Mean ranks of RMSEs for relative risk = 1.5 and different PSUs are sampled for each simulation

| Sampling method | Mean ranks when estimating | | | | | |
|---|---|---|---|---|---|---|
| | Prevalence | | | Relative Risk (RR) | | |
| | *n=7* | *15* | *30* | *n=7* | *15* | *30* |
| SA | 2.70 | 2.76 | 2.64 | 3.30 | 3.54 | 3.34 |
| Quad | 2.38 | 2.48 | 2.54 | 1.66 | 1.56 | 1.96 |
| Square | 1.18 | 1.02 | 1.10 | 2.14 | 1.68 | 1.28 |
| EPI | 3.74 | 3.74 | 3.72 | 2.90 | 3.22 | 3.42 |

See footnote to Table S1

Table S7. Mean ranks of RMSEs for relative risk = 2.0 and different PSUs are sampled for each simulation

| Sampling method | Mean ranks when estimating | | | | | |
|---|---|---|---|---|---|---|
| | Prevalence | | | Relative Risk (RR) | | |
| | *n=7* | *15* | *30* | *n=7* | *15* | *30* |
| SA | 2.70 | 2.80 | 2.76 | 3.30 | 3.64 | 3.38 |
| Quad | 2.34 | 2.38 | 2.44 | 1.52 | 1.50 | 1.98 |
| Square | 1.18 | 1.04 | 1.10 | 2.26 | 1.66 | 1.28 |
| EPI | 3.78 | 3.78 | 3.70 | 2.92 | 3.20 | 3.36 |

See footnote to Table S1

Table S8. Mean ranks of RMSEs for relative risk = 3.0 and different PSUs are sampled for each simulation

| Sampling method | Mean ranks when estimating | | | | | |
|---|---|---|---|---|---|---|
| | Prevalence | | | Relative Risk (RR) | | |
| | *n=7* | *15* | *30* | *n=7* | *15* | *30* |
| SA | 2.56 | 2.74 | 2.64 | 3.54 | 3.46 | 3.20 |
| Quad | 2.40 | 2.36 | 2.42 | 1.38 | 1.52 | 1.98 |
| Square | 1.18 | 1.08 | 1.14 | 2.20 | 1.76 | 1.30 |
| EPI | 3.86 | 3.82 | 3.80 | 2.88 | 3.26 | 3.52 |

See footnote to Table S1

**Tables of Mean Ratios of MSEs**

Table S9. Mean ratios of RMSEs for relative risk = 1.0 and same PSUs are sampled

| Sampling method | Mean ratios when estimating | | | | | |
|---|---|---|---|---|---|---|
| | Prevalence | | | Relative Risk (RR) | | |
| | *n=7* | *15* | *30* | *n=7* | *15* | *30* |
| SA | 1.18 | 1.41 | 1.62 | 1.08 | 1.31 | 1.44 |
| Quad | 1.23 | 1.43 | 1.75 | 1.33 | 1.05 | 1.16 |
| Square | 1.01 | 1.04 | 1.07 | 1.22 | 1.00 | 0.99 |
| EPI | 1.39 | 1.73 | 2.15 | 1.49 | 1.34 | 1.55 |

Note: RMSE = Root Mean Squared Error. PSU = Primary Sampling Unit. The first three columns of data show the mean RMSE ratios (ratio of RMSE for the sampling method: RMSE for simple random sampling) for the prevalence estimates for the three sample sizes within clusters (n=7, 15, or 30). The last three columns show the mean RMSE ratios for estimates of relative risks.

Table S10. Mean ratios of RMSEs for relative risk = 1.5 and same PSUs are sampled

| Sampling method | Mean ratios when estimating | | | | | |
|---|---|---|---|---|---|---|
| | Prevalence | | | Relative Risk (RR) | | |
| | *n=7* | *15* | *30* | *n=7* | *15* | *30* |
| SA | 1.22 | 1.48 | 1.81 | 1.09 | 1.27 | 1.39 |
| Quad | 1.27 | 1.50 | 1.86 | 0.98 | 1.03 | 1.17 |
| Square | 1.02 | 1.04 | 1.08 | 0.97 | 0.99 | 1.01 |
| EPI | 1.45 | 1.81 | 2.31 | 1.14 | 1.30 | 1.56 |

See footnote to Table S9.

Table S11. Mean ratios of RMSEs for relative risk = 2.0 and same PSUs are sampled

| Sampling method | Mean ratios when estimating | | | | | |
|---|---|---|---|---|---|---|
| | Prevalence | | | Relative Risk (RR) | | |
| | *n=7* | *15* | *30* | *n=7* | *15* | *30* |
| SA | 1.23 | 1.53 | 1.82 | 1.09 | 1.31 | 1.45 |
| Quad | 1.29 | 1.55 | 1.92 | 0.98 | 1.04 | 1.14 |
| Square | 1.02 | 1.06 | 1.09 | 0.99 | 1.00 | 1.00 |
| EPI | 1.47 | 1.88 | 2.38 | 1.14 | 1.32 | 1.55 |

See footnote to Table S9.

Table S12. Mean ratios of RMSEs for relative risk = 3.0 and same PSUs are sampled

| Sampling method | Mean ratios when estimating Prevalence | | | Relative Risk (RR) | | |
|---|---|---|---|---|---|---|
| | *n=7* | *15* | *30* | *n=7* | *15* | *30* |
| SA | 1.26 | 1.55 | 1.83 | 1.15 | 1.32 | 1.41 |
| Quad | 1.35 | 1.63 | 2.01 | 1.00 | 1.03 | 1.20 |
| Square | 1.03 | 1.06 | 1.11 | 1.00 | 0.98 | 0.99 |
| EPI | 1.52 | 1.94 | 2.43 | 1.16 | 1.32 | 1.58 |

See footnote to Table S9.

Table S13. Mean ratios of RMSEs for relative risk = 1.0 and different PSUs are sampled

| Sampling method | Mean ratios when estimating Prevalence | | | Relative Risk (RR) | | |
|---|---|---|---|---|---|---|
| | *n=7* | *15* | *30* | *n=7* | *15* | *30* |
| SA | 1.19 | 1.42 | 1.60 | 1.09 | 1.27 | 1.35 |
| Quad | 1.24 | 1.43 | 1.75 | 1.62 | 1.00 | 1.10 |
| Square | 1.02 | 1.03 | 1.05 | 0.98 | 1.00 | 1.01 |
| EPI | 1.41 | 1.73 | 2.15 | 1.72 | 1.24 | 1.47 |

See footnote to Table S9.

Table S14. Mean ratios of RMSEs for relative risk = 1.5 and different PSUs are sampled

| Sampling method | Mean ratios when estimating Prevalence | | | Relative Risk (RR) | | |
|---|---|---|---|---|---|---|
| | *n=7* | *15* | *30* | *n=7* | *15* | *30* |
| SA | 1.22 | 1.49 | 1.71 | 1.11 | 1.28 | 1.40 |
| Quad | 1.28 | 1.50 | 1.85 | 0.95 | 0.99 | 1.11 |
| Square | 1.02 | 1.04 | 1.07 | 0.99 | 1.00 | 1.00 |
| EPI | 1.46 | 1.83 | 2.29 | 1.07 | 1.24 | 1.48 |

See footnote to Table S9.

Table S15. Mean ratios of RMSEs for relative risk = 2.0 and different PSUs are sampled

| Sampling method | Mean ratios when estimating | | | | | |
|---|---|---|---|---|---|---|
| | Prevalence | | | Relative Risk (RR) | | |
| | *n=7* | *15* | *30* | *n=7* | *15* | *30* |
| SA | 1.23 | 1.52 | 1.79 | 1.12 | 1.30 | 1.40 |
| Quad | 1.30 | 1.54 | 1.89 | 0.94 | 0.99 | 1.12 |
| Square | 1.02 | 1.05 | 1.07 | 1.00 | 1.00 | 0.99 |
| EPI | 1.49 | 1.88 | 2.33 | 1.06 | 1.25 | 1.47 |

See footnote to Table S9.

Table S16. Mean ratios of RMSEs for relative risk = 3.0 and different PSUs are sampled

| Sampling method | Mean ratios when estimating | | | | | |
|---|---|---|---|---|---|---|
| | Prevalence | | | Relative Risk (RR) | | |
| | *n=7* | *15* | *30* | *n=7* | *15* | *30* |
| SA | 1.25 | 1.53 | 1.71 | 1.14 | 1.28 | 1.35 |
| Quad | 1.34 | 1.60 | 1.91 | 0.94 | 0.99 | 1.11 |
| Square | 1.03 | 1.05 | 1.08 | 1.01 | 1.01 | 1.00 |
| EPI | 1.53 | 1.90 | 2.31 | 1.08 | 1.27 | 1.51 |

See footnote to Table S9.

**Tables of results with eight sampling methods**

**Tables of Mean Ranks**

Table S17. Mean ranks of RMSEs for Relative Risk = 1.0 and same towns are sampled

| Sampling method | Mean ranks when estimating | | | | | |
|---|---|---|---|---|---|---|
| | Prevalence | | | Relative Risk (RR) | | |
| | *n=7* | *15* | *30* | *n=7* | *15* | *30* |
| SA | 4.2 | 4.2 | 4.6 | 5.1 | 5.5 | 5.5 |
| Grid | 5.8 | 6.1 | 6.0 | 6.4 | 6.8 | 7.2 |
| Peri | 3.7 | 4.0 | 3.9 | 3.3 | 3.9 | 3.9 |
| Quad | 3.4 | 3.2 | 3.6 | 2.3 | 2.2 | 2.2 |
| Square | 1.5 | 1.2 | 1.3 | 3.5 | 2.3 | 1.8 |
| EPI | 6.7 | 7.1 | 7.0 | 5.5 | 5.7 | 6.2 |
| EPI3 | 5.9 | 5.7 | 5.4 | 4.8 | 4.8 | 4.8 |
| EPI5 | 4.8 | 4.5 | 4.2 | 5.1 | 4.8 | 4.3 |

See footnote to Table S1

Table S18. Mean ranks of the MSEs when the Relative risk is 1.0 and different towns are sampled

| Sampling method | Mean ranks when estimating | | | | | |
|---|---|---|---|---|---|---|
| | Prevalence | | | Relative Risk (RR) | | |
| | *n=7* | *15* | *30* | *n=7* | *15* | *30* |
| SA | 3.7 | 4.2 | 4.3 | 5.8 | 5.8 | 5.8 |
| Grid | 5.8 | 6.1 | 6.0 | 6.2 | 6.8 | 7.2 |
| Peri | 3.9 | 3.9 | 4.1 | 3.5 | 4.0 | 3.8 |
| Quad | 3.3 | 3.6 | 3.9 | 2.3 | 2.1 | 2.3 |
| Square | 1.5 | 1.2 | 1.3 | 3.6 | 2.3 | 1.9 |
| EPI | 6.8 | 7.1 | 7.0 | 5.5 | 5.7 | 6.2 |
| EPI3 | 5.6 | 5.3 | 5.1 | 4.5 | 4.8 | 5.0 |
| EPI5 | 5.3 | 4.5 | 4.2 | 4.6 | 4.4 | 3.9 |

See footnote to Table S1

Table S19. Mean ranks of RMSEs for Relative Risk = 1.5 and same towns are sampled

| Sampling method | Mean ranks when estimating Prevalence | | | Relative Risk (RR) | | |
|---|---|---|---|---|---|---|
| | *n=7* | *15* | *30* | *n=7* | *15* | *30* |
| SA | 4.0 | 4.2 | 4.5 | 4.8 | 5.6 | 5.6 |
| Grid | 6.0 | 6.1 | 6.2 | 6.6 | 7.1 | 7.0 |
| Peri | 3.6 | 3.5 | 3.6 | 3.5 | 3.6 | 3.8 |
| Quad | 3.4 | 3.5 | 3.7 | 2.6 | 2.1 | 2.5 |
| Square | 1.6 | 1.4 | 1.3 | 3.2 | 2.2 | 1.8 |
| EPI | 6.5 | 7.1 | 6.9 | 5.4 | 5.8 | 6.3 |
| EPI3 | 5.8 | 5.7 | 5.5 | 4.7 | 4.9 | 4.8 |
| EPI5 | 5.0 | 4.6 | 4.2 | 5.2 | 4.6 | 4.2 |

See footnote to Table S1

Table S20. Mean ranks of the MSEs when the Relative risk is 1.5 and different towns are sampled

| Sampling method | Mean ranks when estimating Prevalence | | | Relative Risk (RR) | | |
|---|---|---|---|---|---|---|
| | *n=7* | *15* | *30* | *n=7* | *15* | *30* |
| SA | 4.0 | 4.2 | 4.3 | 6.0 | 6.4 | 5.9 |
| Grid | 5.6 | 5.9 | 5.9 | 6.4 | 6.8 | 6.9 |
| Peri | 3.7 | 3.5 | 3.7 | 3.2 | 3.3 | 3.8 |
| Quad | 3.5 | 3.7 | 4.0 | 2.7 | 1.9 | 2.4 |
| Square | 1.5 | 1.4 | 1.5 | 3.9 | 2.5 | 1.6 |
| EPI | 6.8 | 7.1 | 7.1 | 5.2 | 5.5 | 6.3 |
| EPI3 | 5.6 | 5.5 | 5.3 | 4.4 | 5.1 | 5.0 |
| EPI5 | 5.1 | 4.7 | 4.2 | 4.2 | 4.4 | 4.1 |

See footnote to Table S1

Table S21. Mean ranks of RMSEs for Relative Risk = 2.0 and same towns are sampled

| Sampling method | Mean ranks when estimating Prevalence | | | Relative Risk (RR) | | |
|---|---|---|---|---|---|---|
| | *n=7* | *15* | *30* | *n=7* | *15* | *30* |
| SA | 4.0 | 4.3 | 4.6 | 5.2 | 5.6 | 5.7 |
| Grid | 5.9 | 5.9 | 6.1 | 6.7 | 7.0 | 7.1 |
| Peri | 3.3 | 3.7 | 3.7 | 3.5 | 3.7 | 3.5 |
| Quad | 3.5 | 3.4 | 3.7 | 2.8 | 2.5 | 2.4 |
| Square | 1.5 | 1.3 | 1.2 | 3.0 | 2.3 | 2.0 |
| EPI | 6.7 | 7.0 | 7.1 | 5.4 | 5.6 | 6.4 |
| EPI3 | 5.7 | 5.6 | 5.5 | 4.8 | 4.9 | 4.9 |
| EPI5 | 5.4 | 4.8 | 4.2 | 4.6 | 4.4 | 4.1 |

See footnote to Table S1

Table S22. Mean ranks of the MSEs when the Relative risk is 2.0 and different towns are sampled

| Sampling method | Mean ranks when estimating Prevalence | | | Relative Risk (RR) | | |
|---|---|---|---|---|---|---|
| | *n=7* | *15* | *30* | *n=7* | *15* | *30* |
| SA | 4.0 | 4.4 | 4.4 | 5.8 | 6.3 | 6.0 |
| Grid | 5.8 | 5.8 | 6.0 | 6.8 | 6.7 | 7.2 |
| Peri | 3.3 | 3.6 | 3.5 | 2.9 | 3.6 | 3.7 |
| Quad | 3.5 | 3.7 | 3.9 | 2.4 | 2.0 | 2.3 |
| Square | 1.6 | 1.1 | 1.5 | 3.6 | 2.3 | 1.7 |
| EPI | 6.8 | 6.9 | 7.0 | 5.1 | 5.5 | 6.2 |
| EPI3 | 5.8 | 5.8 | 5.4 | 4.7 | 4.8 | 4.9 |
| EPI5 | 5.1 | 4.7 | 4.2 | 4.7 | 4.8 | 4.0 |

See footnote to Table S1

Table S23. Mean ranks of RMSEs for Relative Risk = 3.0 and same towns are sampled

| Sampling method | Mean ranks when estimating Prevalence | | | Relative Risk (RR) | | |
|---|---|---|---|---|---|---|
| | *n=7* | *15* | *30* | *n=7* | *15* | *30* |
| SA | 4.2 | 4.4 | 4.3 | 5.5 | 5.9 | 5.5 |
| Grid | 5.4 | 5.7 | 5.8 | 6.5 | 6.7 | 6.9 |
| Peri | 3.4 | 3.4 | 3.3 | 3.1 | 3.5 | 3.7 |
| Quad | 3.7 | 3.5 | 4.0 | 2.6 | 1.9 | 2.7 |
| Square | 1.7 | 1.5 | 1.6 | 3.2 | 2.4 | 1.7 |
| EPI | 6.7 | 6.9 | 7.1 | 5.7 | 5.8 | 6.4 |
| EPI3 | 5.7 | 5.7 | 5.5 | 4.8 | 5.1 | 5.0 |
| EPI5 | 5.1 | 4.8 | 4.5 | 4.6 | 4.7 | 4.0 |

See footnote to Table S1

Table S24. Mean ranks of the MSEs when the Relative risk is 3.0 and different towns are sampled

| Sampling method | Mean ranks when estimating Prevalence | | | Relative Risk (RR) | | |
|---|---|---|---|---|---|---|
| | *n=7* | *15* | *30* | *n=7* | *15* | *30* |
| SA | 3.8 | 4.5 | 4.2 | 6.2 | 6.0 | 5.6 |
| Grid | 5.4 | 5.8 | 6.0 | 6.4 | 7.2 | 7.2 |
| Peri | 3.5 | 3.5 | 3.3 | 2.9 | 3.0 | 3.5 |
| Quad | 3.6 | 3.5 | 4.0 | 2.3 | 1.9 | 2.4 |
| Square | 1.5 | 1.2 | 1.4 | 3.8 | 2.8 | 1.7 |
| EPI | 7.1 | 7.1 | 7.3 | 4.9 | 5.7 | 6.5 |
| EPI3 | 6.2 | 5.8 | 5.5 | 5.0 | 5.2 | 5.1 |
| EPI5 | 5.0 | 4.6 | 4.3 | 4.5 | 4.2 | 4.2 |

See footnote to Table S1

**Tables of Mean Ratios of MSEs**

Table S25. Mean ratios of RMSEs for Relative Risk = 1.0 and same towns are sampled

| Sampling method | Mean ratios when estimating Prevalence | | | Relative Risk (RR) | | |
|---|---|---|---|---|---|---|
| | *n=7* | *15* | *30* | *n=7* | *15* | *30* |
| SA | 1.18 | 1.41 | 1.62 | 1.08 | 1.31 | 1.44 |
| Grid | 1.28 | 1.55 | 1.92 | 1.35 | 1.46 | 1.68 |
| Peri | 1.19 | 1.43 | 1.74 | 1.25 | 1.20 | 1.35 |
| Quad | 1.23 | 1.43 | 1.75 | 1.33 | 1.05 | 1.16 |
| Square | 1.01 | 1.04 | 1.07 | 1.22 | 1.00 | 0.99 |
| EPI | 1.39 | 1.73 | 2.15 | 1.49 | 1.34 | 1.55 |
| EPI3 | 1.37 | 1.64 | 1.97 | 1.13 | 1.28 | 1.44 |
| EPI5 | 1.33 | 1.57 | 1.85 | 1.45 | 1.27 | 1.38 |

See footnote to Table S9

Table S26. Mean ratios of RMSEs for Relative Risk = 1.0 and different towns are sampled

| Sampling method | Mean ratios when estimating Prevalence | | | Relative Risk (RR) | | |
|---|---|---|---|---|---|---|
| | *n=7* | *15* | *30* | *n=7* | *15* | *30* |
| SA | 1.40 | 1.99 | 2.83 | 1.22 | 1.59 | 1.99 |
| Grid | 1.61 | 2.34 | 3.59 | 1.31 | 1.78 | 2.44 |
| Peri | 1.51 | 2.08 | 3.13 | 1.88 | 1.29 | 1.58 |
| Quad | 1.60 | 2.21 | 3.42 | 24.57 | 1.02 | 1.21 |
| Square | 1.05 | 1.05 | 1.11 | 0.99 | 1.01 | 1.02 |
| EPI | 2.03 | 3.13 | 4.91 | 22.76 | 1.57 | 2.18 |
| EPI3 | 1.92 | 2.75 | 4.05 | 24.73 | 1.46 | 1.86 |
| EPI5 | 1.85 | 2.57 | 3.67 | 26.12 | 1.39 | 1.64 |

See footnote to Table S9

Table S27. Mean ratios of RMSEs for Relative Risk = 1.5 and same towns are sampled

| Sampling method | Mean ratios when estimating | | | | | |
|---|---|---|---|---|---|---|
| | Prevalence | | | Relative Risk (RR) | | |
| | *n=7* | *15* | *30* | *n=7* | *15* | *30* |
| SA | 1.47 | 2.17 | 3.67 | 1.23 | 1.63 | 2.19 |
| Grid | 1.72 | 2.62 | 4.22 | 1.44 | 2.14 | 2.83 |
| Peri | 1.56 | 2.21 | 3.44 | 1.11 | 1.31 | 1.72 |
| Quad | 1.68 | 2.47 | 3.93 | 0.99 | 1.08 | 1.40 |
| Square | 1.04 | 1.08 | 1.17 | 0.97 | 0.99 | 1.01 |
| EPI | 2.15 | 3.46 | 5.74 | 1.36 | 1.73 | 2.54 |
| EPI3 | 2.06 | 3.13 | 4.83 | 1.27 | 1.61 | 2.17 |
| EPI5 | 1.98 | 2.93 | 4.37 | 1.26 | 1.49 | 1.97 |

See footnote to Table S9

Table S28. Mean ratios of RMSEs for Relative Risk = 1.5 and different towns are sampled

| Sampling method | Mean ratios when estimating | | | | | |
|---|---|---|---|---|---|---|
| | Prevalence | | | Relative Risk (RR) | | |
| | *n=7* | *15* | *30* | *n=7* | *15* | *30* |
| SA | 1.47 | 2.19 | 3.19 | 1.24 | 1.63 | 2/19 |
| Grid | 1.71 | 2.59 | 4.04 | 1.30 | 1.84 | 2.69 |
| Peri | 1.59 | 2.22 | 3.38 | 0.95 | 1.20 | 1.54 |
| Quad | 1.71 | 2.44 | 3.87 | 0.91 | 1.00 | 1.24 |
| Square | 1.04 | 1.08 | 1.15 | 0.99 | 1.01 | 1.02 |
| EPI | 2.19 | 3.48 | 5.60 | 1.15 | 1.54 | 2.20 |
| EPI3 | 2.08 | 3.07 | 4.62 | 1.06 | 1.46 | 1.90 |
| EPI5 | 1.99 | 2.85 | 4.19 | 1.08 | 1.35 | 1.72 |

See footnote to Table S9

Table S29. Mean ratios of RMSEs for Relative Risk = 2.0 and same towns are sampled

| Sampling method | Mean ratios when estimating Prevalence | | | Relative Risk (RR) | | |
|---|---|---|---|---|---|---|
| | *n=7* | *15* | *30* | *n=7* | *15* | *30* |
| SA | 1.50 | 2.32 | 3.63 | 1.22 | 1.72 | 2.40 |
| Grid | 1.75 | 2.76 | 4.38 | 1.60 | 2.09 | 2.77 |
| Peri | 1.48 | 2.22 | 3.53 | 1.08 | 1.32 | 1.66 |
| Quad | 1.74 | 2.61 | 4.18 | 1.01 | 1.10 | 1.35 |
| Square | 1.04 | 1.12 | 1.20 | 0.99 | 1.01 | 1.00 |
| EPI | 2.23 | 3.71 | 6.03 | 1.34 | 1.78 | 2.48 |
| EPI3 | 2.13 | 3.35 | 5.15 | 1.29 | 1.59 | 2.09 |
| EPI5 | 2.08 | 3.18 | 4.64 | 1.25 | 1.48 | 1.84 |

See footnote to Table S9

Table S30. Mean ratios of RMSEs for Relative Risk = 2.0 and different towns are sampled

| Sampling method | Mean ratios when estimating Prevalence | | | Relative Risk (RR) | | |
|---|---|---|---|---|---|---|
| | *n=7* | *15* | *30* | *n=7* | *15* | *30* |
| SA | 1.50 | 2.27 | 3.44 | 1.28 | 1.69 | 2.16 |
| Grid | 1.74 | 2.68 | 4.13 | 1.44 | 1.88 | 2.50 |
| Peri | 1.54 | 2.18 | 3.31 | 0.96 | 1.25 | 1.55 |
| Quad | 1.77 | 2.57 | 3.95 | 0.92 | 0.99 | 1.25 |
| Square | 1.04 | 1.10 | 1.16 | 1.00 | 1.00 | 0.99 |
| EPI | 2.27 | 3.65 | 5.70 | 1.16 | 1.59 | 2.15 |
| EPI3 | 2.15 | 3.24 | 4.72 | 1.15 | 1.45 | 1.84 |
| EPI5 | 2.06 | 2.99 | 4.31 | 1.13 | 1.40 | 1.68 |

See footnote to Table S9

Table S31. Mean ratios of RMSEs for Relative Risk = 3.0 and same towns are sampled

| Sampling method | Mean ratios when estimating Prevalence | | | Relative Risk (RR) | | |
|---|---|---|---|---|---|---|
| | *n=7* | *15* | *30* | *n=7* | *15* | *30* |
| SA | 1.26 | 1.55 | 1.83 | 1.15 | 1.32 | 1.41 |
| Grid | 1.35 | 1.69 | 2.12 | 1.23 | 1.45 | 1.67 |
| Peri | 1.23 | 1.48 | 1.81 | 1.00 | 1.13 | 1.32 |
| Quad | 1.35 | 1.63 | 2.01 | 1.00 | 1.03 | 1.20 |
| Square | 1.03 | 1.06 | 1.11 | 1.00 | 0.98 | 0.99 |
| EPI | 1.52 | 1.94 | 2.43 | 1.16 | 1.32 | 1.58 |
| EPI3 | 1.49 | 1.84 | 2.24 | 1.12 | 1.27 | 1.45 |
| EPI5 | 1.46 | 1.79 | 2.14 | 1.11 | 1.26 | 1.36 |

See footnote to Table S9

Table S32. Mean ratios of RMSEs for Relative Risk = 3.0 and different towns are sampled

| Sampling method | Mean ratios when estimating Prevalence | | | Relative Risk (RR) | | |
|---|---|---|---|---|---|---|
| | *n=7* | *15* | *30* | *n=7* | *15* | *30* |
| SA | 1.55 | 2.30 | 3.16 | 1.30 | 1.63 | 2.01 |
| Grid | 1.77 | 2.67 | 3.94 | 1.39 | 1.89 | 2.63 |
| Peri | 1.58 | 2.10 | 2.99 | 0.95 | 1.18 | 1.56 |
| Quad | 1.87 | 2.70 | 3.99 | 0.90 | 1.00 | 1.26 |
| Square | 1.06 | 1.11 | 1.17 | 1.03 | 1.03 | 1.01 |
| EPI | 2.38 | 3.68 | 5.52 | 1.18 | 1.63 | 2.30 |
| EPI3 | 2.25 | 3.30 | 4.62 | 1.16 | 1.53 | 1.91 |
| EPI5 | 2.16 | 3.03 | 4.22 | 1.11 | 1.41 | 1.70 |

See footnote to Table S9

**How to interpret the Excel files of results**

This section will guide readers through the sheets of the Excel files of results. There are two files. The first, *Results4x50*, and presents the results used in the manuscript. The second, *Results8x50*, shows results for all eight sampling methods, which include those not presented in the manuscript. The basic format of both files is the same.

*Parameters*

The first sheet, ‘parameters’, shows possible values for the parameters used.  We note that the program was written to allow the user to include more parameters than we did.  For example, we determined population density based on a linear relationship between the x and y coordinates of the towns.  We could have used a quadratic relationship.

For parameters whose input is ‘Range’, the three values are the minimum value, the maximum value, and the increment size.  The minimum value was not used.  Thus for target prevalence, the minimum value was 0.1, the maximum was 0.5, and the increment was 0.008.  The values used were 1.008, 1.016, …, 0.5.

For the values whose input is ‘array’, the list shows the values which were used.  This was done when it was not possible to use a minimum, maximum and increment value.

To vary the value of certain parameters across the towns in a population, some of parameters are made up of three comma-separated values.  The first number represents the base value used for all towns in the population, the second is the linear regression coefficient, and the third is the residual variance.

Lines 56-58 show the weights given to income, risk, and age in the Disease Determination Algorithm.

*Populations*

This sheet shows the particular combination of parameter values used in each of the 50 populations.

*RR*

The next eight sheets show the results for the four values of Relative Risk, and for when we did or did not use the same towns/clusters for the 1,000 simulated samples. When we used a different set of towns, we label the sheet ‘Town resample’.

The first table in each of these sheets shows the means of the ratios of each sampling method’s MSE to the MSE for Simple Random Sampling. This is done for estimates of Prevalence and RR, and for the three sample sizes per cluster (7, 15, and 30). The Table to the right shows the equivalent results when we first took the square roots of the MSEs and then computed the ratios. These are the values used in Tables in the main document.

The next table repeats this information, along with more details on the ratios – their minimum and maximum values and their standard deviations (SDs).

The next three tables show for each of the sample sizes how often each sampling method’s RMSE was ranked 1, 2, 3, 4 for when we restricted the number of sampling methods being compared to the four in the main document (‘Situation 1’) or ranked 1, 2, …, 8 for when we considered the eight sampling methods noted above (‘Situation 2’). For Situation 1, The first 4 columns (B to E) show the ranks when estimating prevalence, the next 4 (F to I) when estimating RR. For Situation 2, the first 8 columns (B to I) show the ranks when estimating prevalence, the next 8 (J to Q) when estimating RR.

The next 50 tables show results for each population. Columns B and C show the population values of Prevalence and RR, respectively. The next three columns show for n=7 per cluster the mean, SD, and MSE for sample estimates of prevalence; the next four show the mean RR, SD RR, pooled RR (treating the clusters as separate strata), and MSE for estimates of RR. These seven columns are then repeated for the other two sample sizes, n=15 and 30.

The final three tables show the results for the three populations for which there was no variation – every individual had the same probability of disease, with a different probability for each population.
